# The Impact of Short-Form Video Use on Cognitive and Mental Health Outcomes: A Systematic Review

**DOI:** 10.1101/2025.08.27.25334540

**Authors:** Sara Arouch, Dan Edgcumbe, Sally Pezaro, Ksenija da Silva

## Abstract

**Background:** Short-form video (SFV) platforms such as TikTok, Instagram Reels, and YouTube Shorts dominate digital engagement, especially among Generation Z. Their rapid, algorithm-driven, and emotionally charged design raises unique concerns about potential impacts on cognitive functions and mental health. Despite growing public discourse, evidence remains fragmented.

**Methods:** We conducted a systematic review in accordance with the PRISMA and Cochrane guidelines. Eight databases (2014–2024) were searched, including PsycINFO, PubMed, Scopus, and CINAHL. Eligible studies investigated the effects of SFV use on cognitive mechanisms (e.g., attention, executive function, memory, emotional regulation) and mental health outcomes among adults aged 18–29. The risk of bias was assessed using the Joanna Briggs Institute checklist, and the findings were narratively synthesised.

**Results:** Of 519,101 initial records, 17 studies met the inclusion criteria. High-frequency SFV use was consistently associated with attentional disruption, reduced executive functioning, and emotional dysregulation. Several studies reported increased anxiety and compulsive use, though findings were heterogeneous. Self-regulatory capacity emerged as a potential moderator. However, evidence was limited by a reliance on cross-sectional designs, self-report measures, and inconsistent operationalisation of cognitive outcomes.

**Conclusion:** SFV use may adversely affect cognitive control and emotional regulation in Generation Z, contributing to heightened risks for anxiety and compulsive behaviours. Given methodological limitations in the current literature, further longitudinal and experimental research is urgently needed. A qualitative follow-up study will explore users’ lived experiences, informing the development of a tailored assessment tool. These findings hold implications for digital psychiatry, public health guidance, and intervention design in young adult populations.

**Key points:** *What is already known on this topic:*

- Short-form video platforms such as TikTok and Instagram Reels are widely used by Generation Z.
- Concerns have been raised about their potential impact on attention, executive function, and mental health.
- Existing research is fragmented, with inconsistent findings and a lack of longitudinal evidence.

*What this study adds:*

- This is the first systematic review synthesising evidence across cognitive mechanisms and mental health outcomes in relation to SFV use.
- Frequent SFV use is associated with attentional disruption, reduced executive function, emotional dysregulation, and increased anxiety/compulsive use.
- Self-regulatory capacity may moderate these effects, suggesting individual differences in vulnerability.

*How this study might affect research, practice, or policy:*

- Highlights the need for longitudinal and experimental studies to clarify causality.
- Supports development of targeted public health messaging around digital self-regulation for young adults.
- Provides a foundation for psychiatry, psychology, and education sectors to address cognitive and emotional risks linked to SFV platforms.

## 1. Background

### 1.1. Overview

#### SFV, Communication and Mental Health

Social media (SM) platforms have transformed global communication, fostering unprecedented connectivity and digital interaction. With 64.2% of the world’s population actively using SM, its influence on social norms, identity formation, and psychological well-being is undeniable (Kemp, 2024). Among adolescents in the U.S., 95% report daily use, with 35% describing themselves as ‘almost constantly’ online (Pew Research Center, 2024). While SM provides opportunities for social connection, self-expression, and educational engagement, its impact on mental health has raised concerns. Studies suggest associations with anxiety, depression, addiction, and body image dissatisfaction, as well as psychological phenomena such as social comparison and Fear of Missing Out (FoMO) (Huang, 2017; Keles et al., 2020). However, the causal mechanisms of these effects remain inconclusive, necessitating further research into how specific social media formats shape mental health outcomes.

One of the most rapidly evolving and cognitively engaging forms of SM content is short-form videos on platforms such as TikTok, Instagram Reels, and YouTube Shorts. Even Google has recently integrated SFV into its search results, reflecting the growing dominance of this content format. These platforms prioritise high-speed, visually stimulating content that maximises user retention through algorithm-driven autoplay and personalised recommendation systems (Qin et al., 2022). While SM may enhance global connectivity, creativity, and information accessibility, it promotes habitual use, attentional fragmentation, and emotional dysregulation (Twenge, 2019). For instance, recent research indicates that TikTok’s short-form content may lead to problematic usage patterns, potentially resulting in adverse mental health outcomes such as increased anxiety, compulsive behaviours, and mood disturbances (Pasquale et al., 2025). The short-form nature of video content engages key cognitive mechanisms, such as attention and executive functions. It may influence behavioural and cognitive processes, raising questions about its potential psychological and mental health implications. Given their widespread adoption and unique design, SFVs warrant further investigation into their cognitive and mental health effects.

#### iGen’s Unique Context

As digital landscapes continue to evolve, younger generations are disproportionately affected by the mental demands of SM and, consequently, SFV. Among them, Generation Z (Gen Z) has grown up fully immersed in these platforms, which have shaped their developmental trajectories and mental health outcomes. Gen Z, born post-1995, represents the first generation to grow up fully immersed in digital environments. This cohort is characterised by high SM engagement, prolonged parental oversight, and delayed traditional adult milestones (Twenge, 2019). Studies suggest Gen Z experiences rank higher rates of mental health issues compared to previous generations, with increased vulnerabilities to anxiety, depression, addiction, and body image concerns correlating with SM use (e.g., frequent Instagram scrolling associated with appearance comparisons) (Odgers & Jensen, 2020). However, the role of cognitive processes and functions in mediating these outcomes remains underexplored.

#### Defining Cognitive Mechanisms and Impact on Mental Health

Cognitive mechanisms, comprising both cognitive processes and cognitive functions, encompass the mental operations and capacities that allow individuals to perceive, interpret, and respond to information effectively. These mechanisms are foundational to essential activities such as reasoning, decision-making, learning, and the regulation of behaviour (Diamond, 2013). Although they serve distinct roles, the terms *“cognitive processes”* and *“cognitive functions”* are often used interchangeably, which contributes to conceptual confusion. Distinguishing between these terms is crucial for understanding how SFV impacts cognitive engagement and mental health (Miyake & Friedman, 2012).

Cognitive processes refer to the fundamental, moment-to-moment mental activities that underlie specific cognitive tasks. These are dynamic operations through which the brain encodes, stores, retrieves, and manipulates information. Key processes pertinent to this study include attention (the selective allocation of mental resources to relevant stimuli while suppressing distractions), memory (the retention and recall of information), and critical thinking (the ability to analyse data, assess alternatives, and make informed decisions).

In contrast, cognitive functions are broader, integrative abilities that emerge from the coordination of multiple cognitive processes. These functions are typically more stable and geared toward enabling complex, goal-directed behaviours. Relevant examples in this context include executive function (the orchestration of thoughts and behaviours through planning, inhibitory control, and cognitive shifting), cognitive flexibility (the capacity to adapt to changing tasks or demands), and emotional regulation (the ability to monitor, manage, and respond to emotional experiences appropriately).

Given the complex, bidirectional nature of the relationship between SM use and cognitive functioning, as well as its broader implications for mental health, further empirical investigation is warranted.

Building on the distinction between cognitive processes and functions, it is crucial to examine how these mechanisms interact with mental health in the context of SFV use. Concerns about the strain on these mechanisms, particularly in emotionally charged and cognitively demanding digital environments like SFV. Executive functions, for example, are central to regulating thoughts, behaviours, and emotions and may be especially vulnerable to continuous digital stimulation and multitasking demands. Disruptions in these systems can contribute to difficulties in attention, self-regulation, and emotional balance—factors closely tied to mental health. Accordingly, this systematic review aims to critically assess current literature to clarify the links between SFV use, cognitive engagement, and mental health and to identify key gaps in existing research.

### 1.2. Rationale

Building on the conceptual distinction between cognitive processes and functions and their established relevance to mental health, it becomes essential to investigate how increased engagement with SFV content affects these cognitive mechanisms. As SFV platforms increasingly influence the digital experiences of Generation Z, questions around their cognitive and mental impact have gained urgency.

Understanding these effects through a systematic review is vital for contextualising current research, appraising the coherence of theoretical models, and evaluating the methodological consistency across studies. Through a comprehensive synthesis of the literature, this review seeks to map prevailing findings, identify recurring conceptual and empirical patterns, and reveal critical gaps in measurement and design. In doing so, it provides a structured foundation for advancing theoretical insights and guiding future empirical research in this emerging area.

#### Objectives

– To systematically identify, synthesise, and evaluate existing studies examining the impact of social media (SM), with a focus on short-form video platforms, on cognitive mechanisms (i.e., cognitive processes and functions) and mental health.
– To explore emerging themes, conceptual trends, and methodological approaches within the literature.
– To highlight inconsistencies, limitations, and underexplored areas that warrant further research.

## 2. Methods

This systematic review was conducted following the PRISMA (Preferred Reporting Items for Systematic Reviews and Meta-Analyses) guidelines to ensure transparency and replicability (Haddaway, Page, Pritchard, & McGuinness, 2022). Methodological guidance was also drawn from the Cochrane Handbook for Systematic Reviews of Interventions to enhance the rigour of the review process. A structured approach was employed for study selection, risk of bias assessment, and evidence synthesis. Current evidence on the relationship between SFV, cognitive mechanisms, and mental health outcomes in Generation Z was systematically synthesised. In alignment with PRISMA and Cochrane standards, the methodology was informed by a comprehensive search strategy, predefined inclusion and exclusion criteria, and a standardised data extraction procedure to ensure methodological integrity.

### 2.1. Eligibility Criteria

#### Inclusion (PICO)

1. Studies focusing on the impact of SFV use on cognitive processes and mental health.
2. Research involving participants aged between 18 and 29 corresponds to the adult Gen Z demographic.
3. Peer-reviewed articles published in English between 1^st^ January 2014 and 18^th^ November 2024.

#### PICO

**PICO:**
|  |  |
| --- | --- |
| **Population** | Adults - Generation Z (18 to 29 years old) -<br>No history of psychiatric diagnoses,<br>substance abuse or current psycho-<br>pharmacological medication. |
| **Intervention** | SFV use and impact on cognitive<br>mechanisms & mental health. |
| **Comparison** | Different levels or patterns of SFV use (e.g.,<br>high vs. low usage, type of usage) |
| **Outcome** | Impact on cognitive mechanisms and<br>mental health. |

#### Exclusion Criteria

1. Studies not directly examining the effects of cognitive mechanisms or mental health.
2. Research focusing exclusively on older adults or children outside the Gen Z age range.
3. Non-peer-reviewed and grey literature (non-peer-reviewed sources such as reports, dissertations, conference proceedings, and policy papers not published in academic journals).
4. Articles published in languages other than English.

### 2.2. Information Sources and Strategy

#### Databases

To ensure comprehensive coverage of relevant studies, the systematic literature review (SLR) was conducted across multiple electronic databases. These included PsycINFO, PsycARTICLES, MEDLINE, Scopus, Academic Search Complete, PubMed, Cochrane, and CINAHL.

#### Search Strategy

The search was carried out between November 10th and November 18th, 2024, using a structured and systematic approach designed to capture the intersection of SFV use, cognitive mechanisms, and mental health. Specific keywords, Boolean operators, and Medical Subject Headings (MeSH) terms were employed and tailored to each database to ensure both breadth and precision.

A search strategy table was prepared to summarise the methodological approach, including the databases searched and the application of search strings and filters. Boolean operators, MeSH terms, and keyword combinations were adapted for each platform to target studies focused on the six cognitive mechanisms under investigation and the Generation Z population.

The use of both standardised MeSH terms and free-text keywords expanded the search’s reach and enabled the identification of studies using varied terminologies. However, due to the broad scope of the search, which encompasses multiple databases and a diverse range of mechanisms, a large number of duplicate records were retrieved in the initial results. As a result, a substantial preliminary dataset was generated. A rigorous, multi-stage screening process was then applied to progressively refine the dataset, ensuring that only the most relevant and methodologically sound studies were included in the final synthesis.

**Figure 1.**
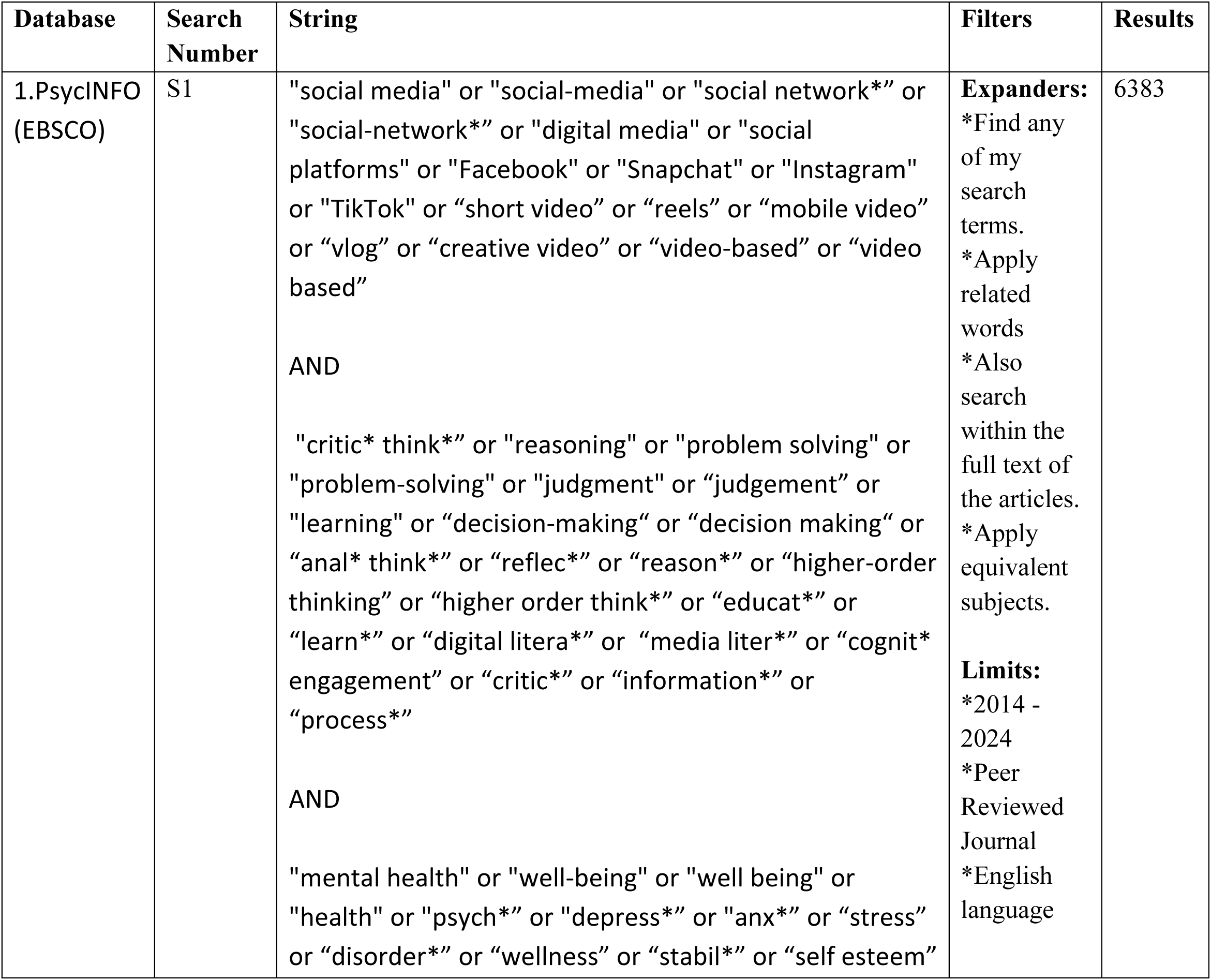
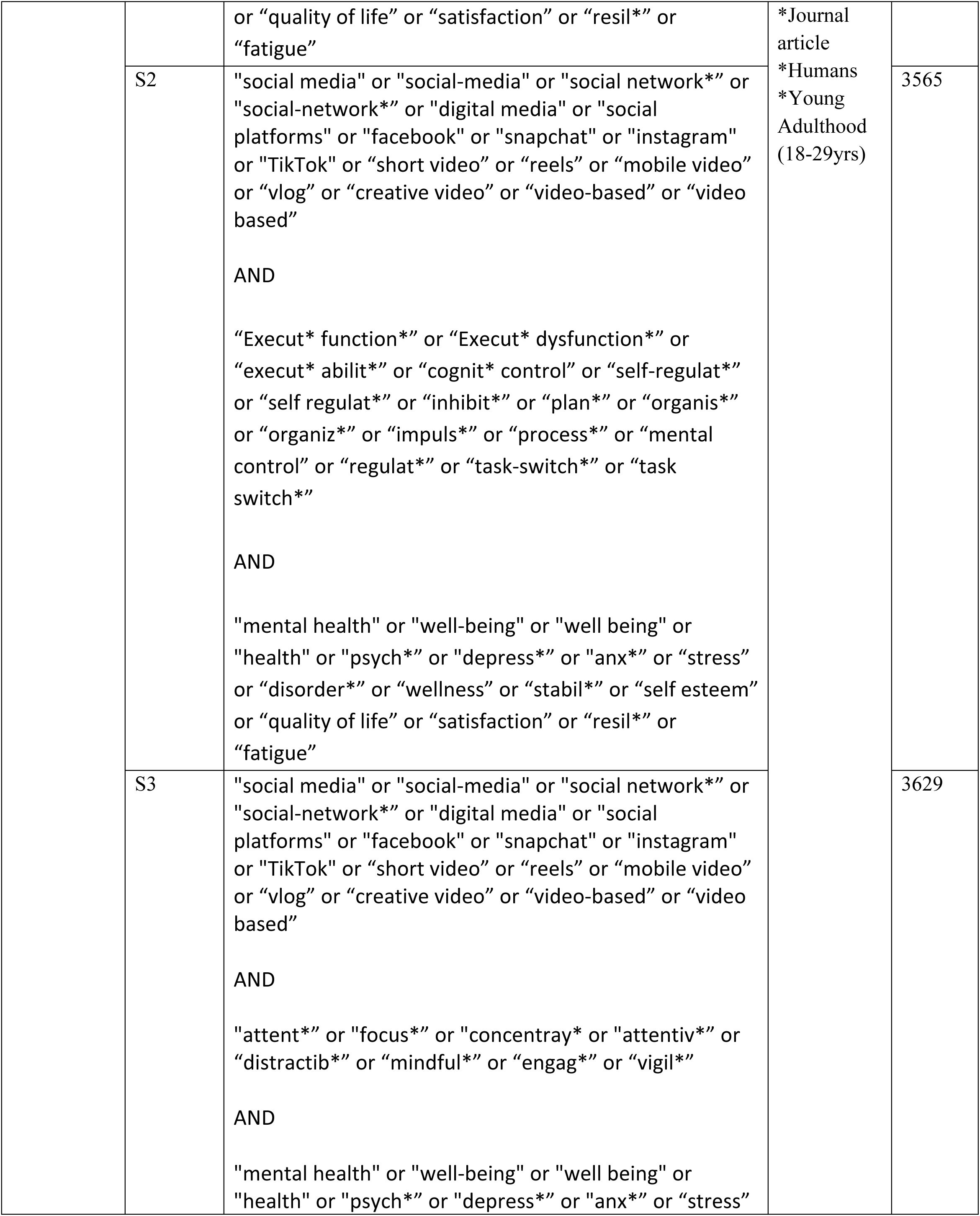
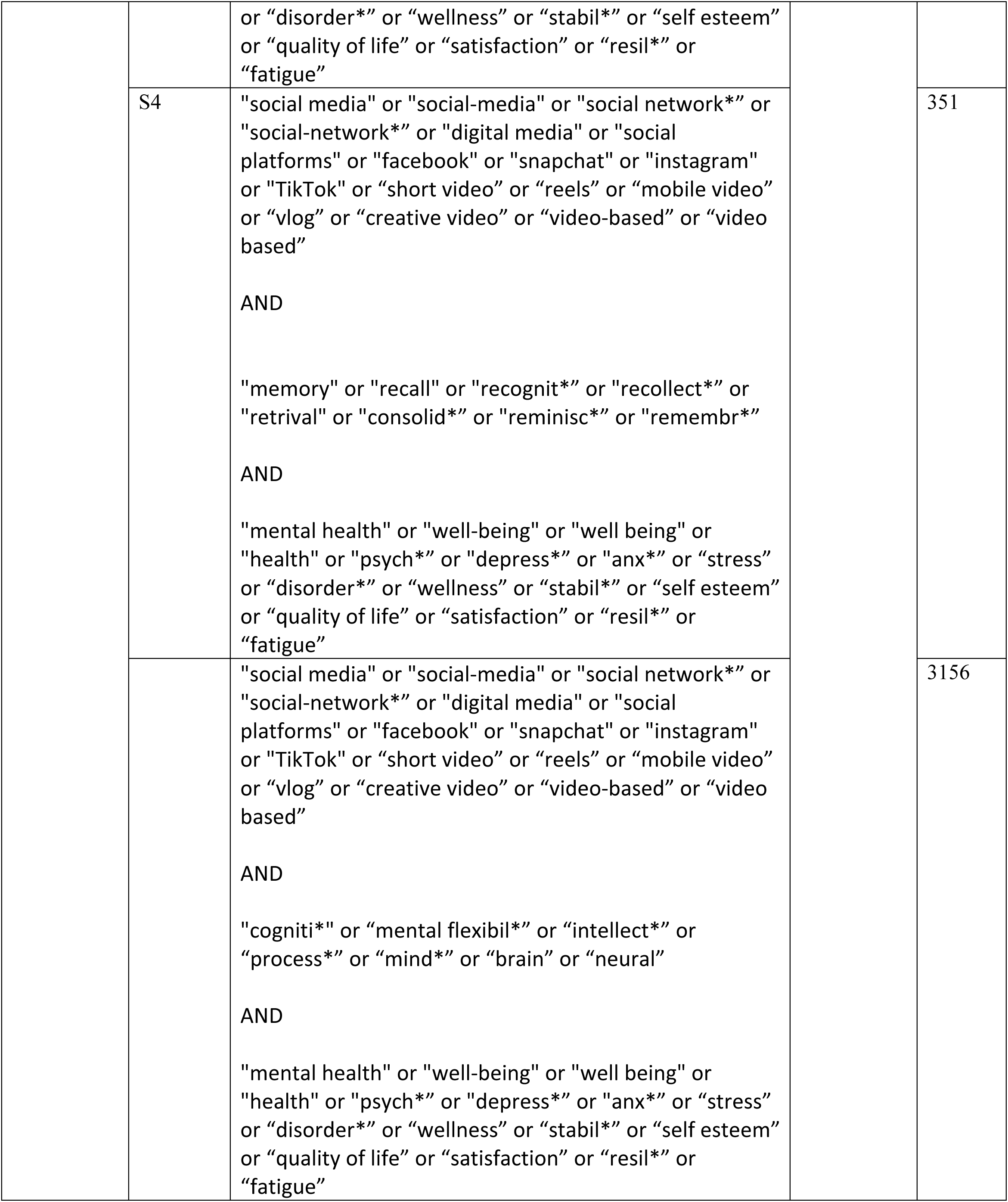
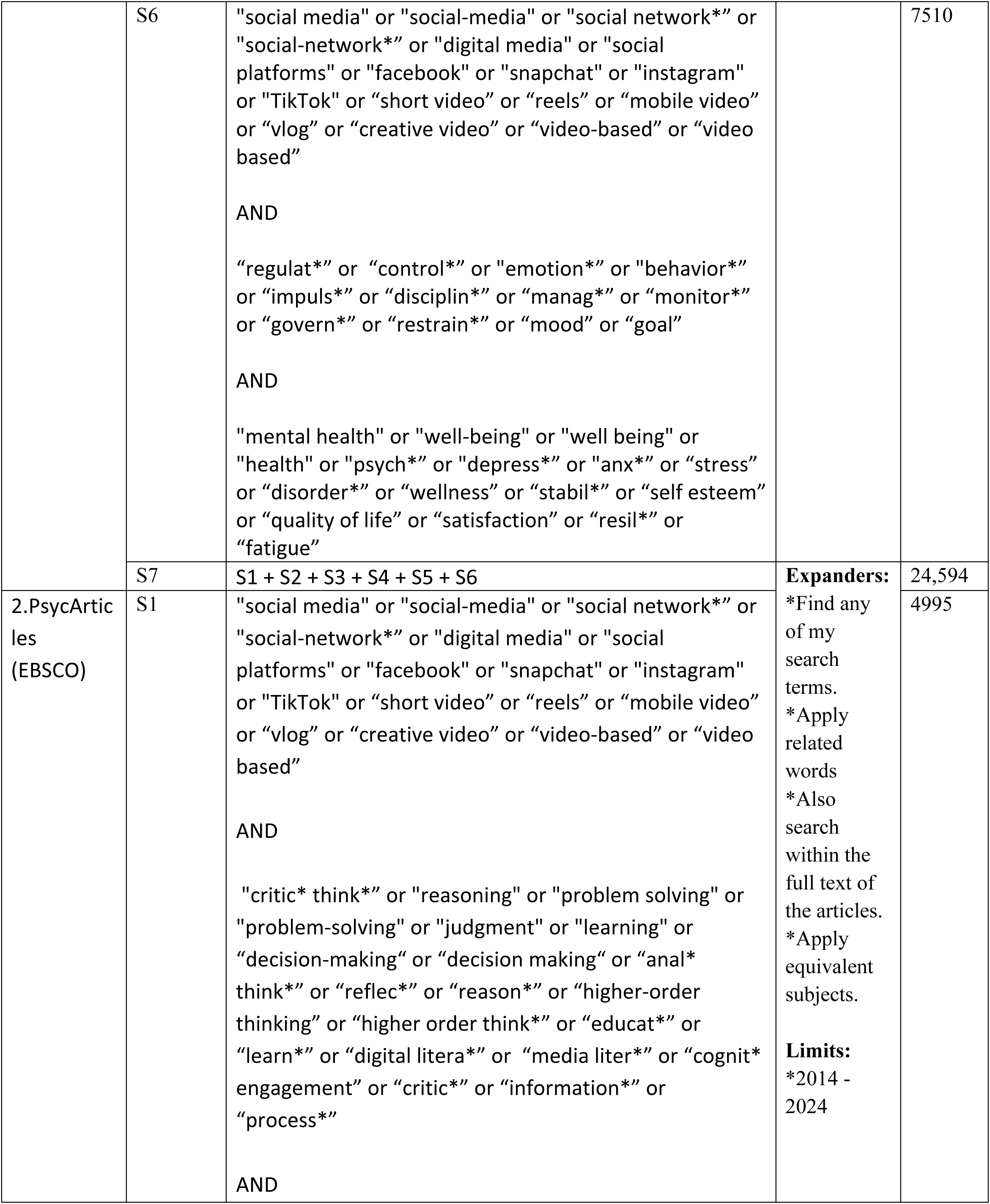
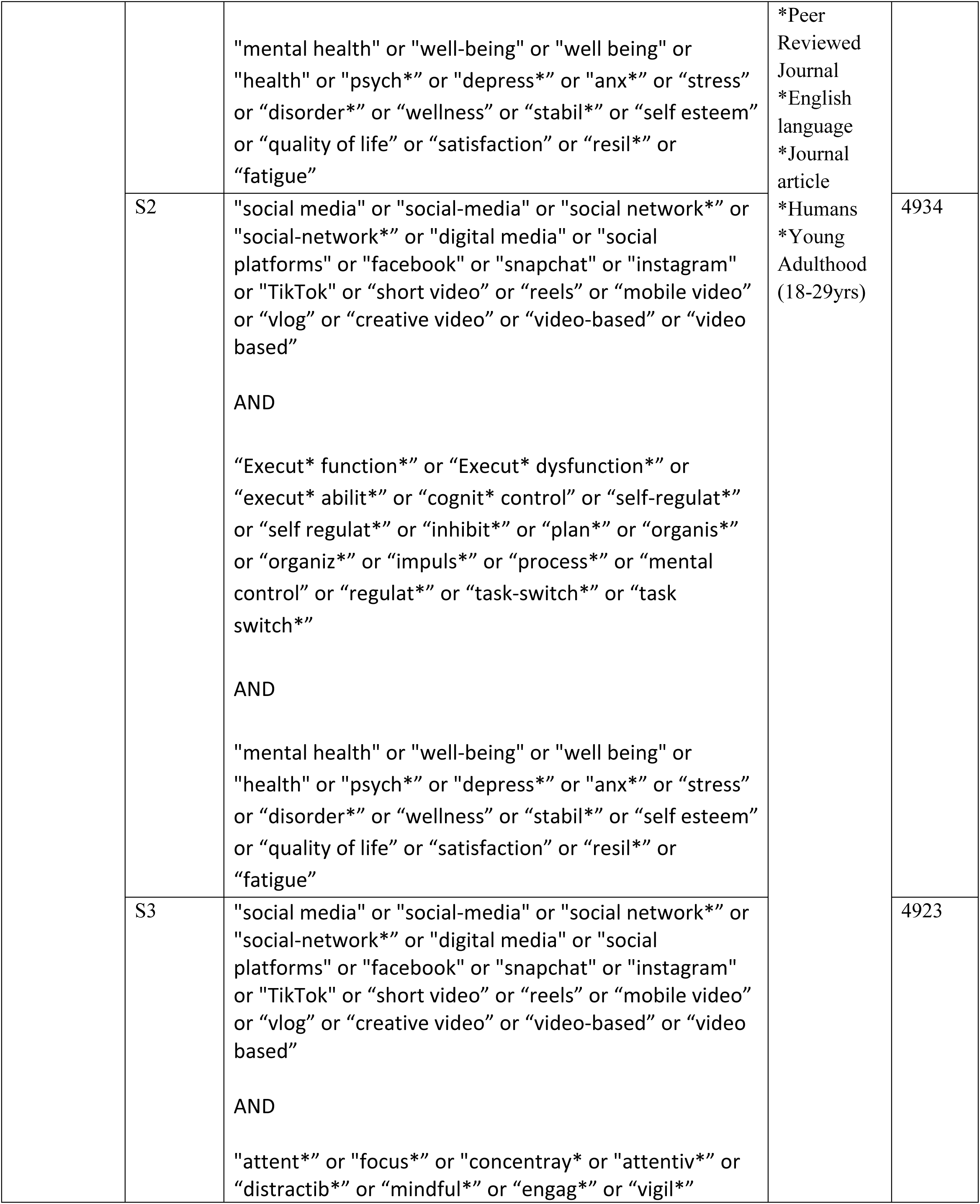
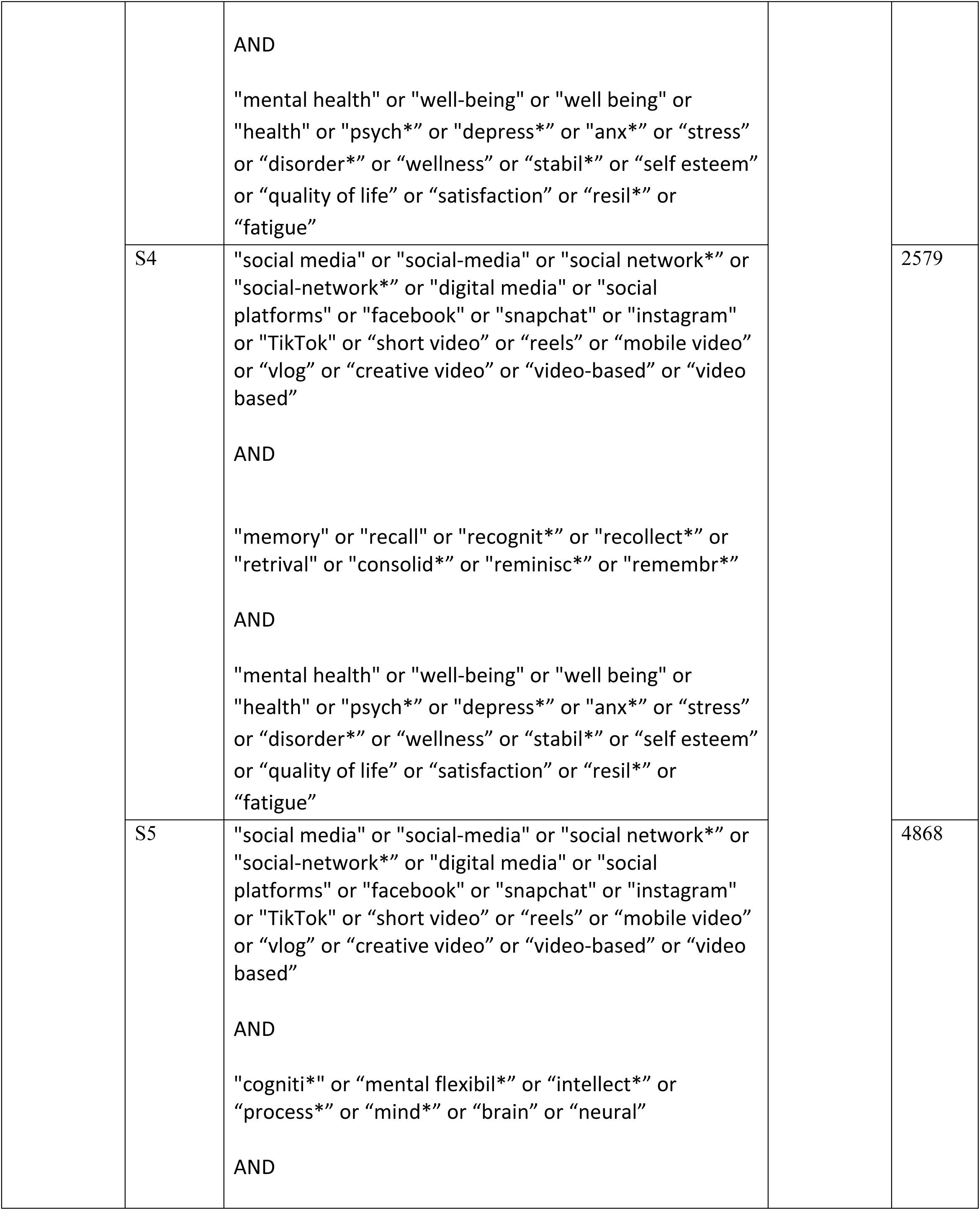
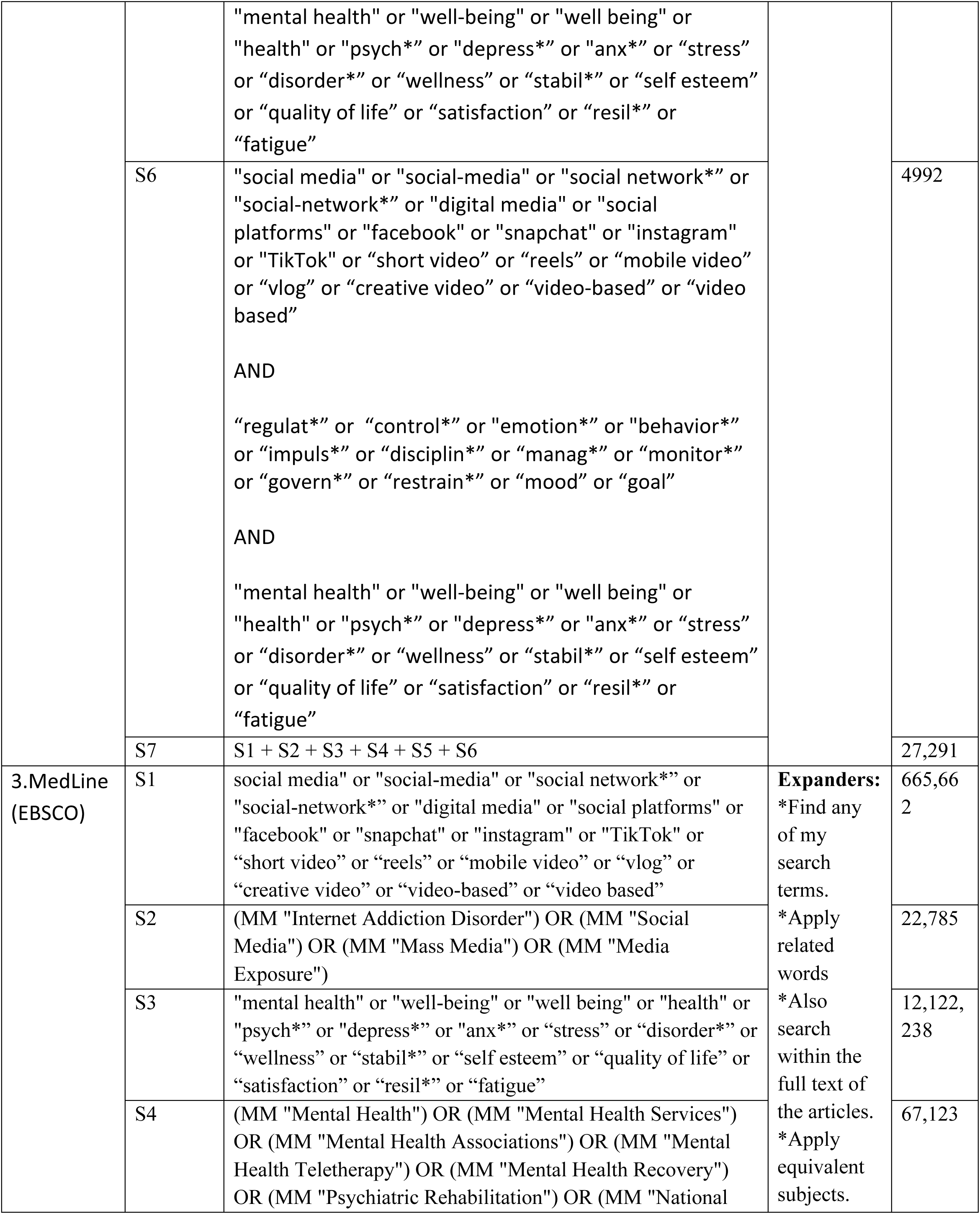
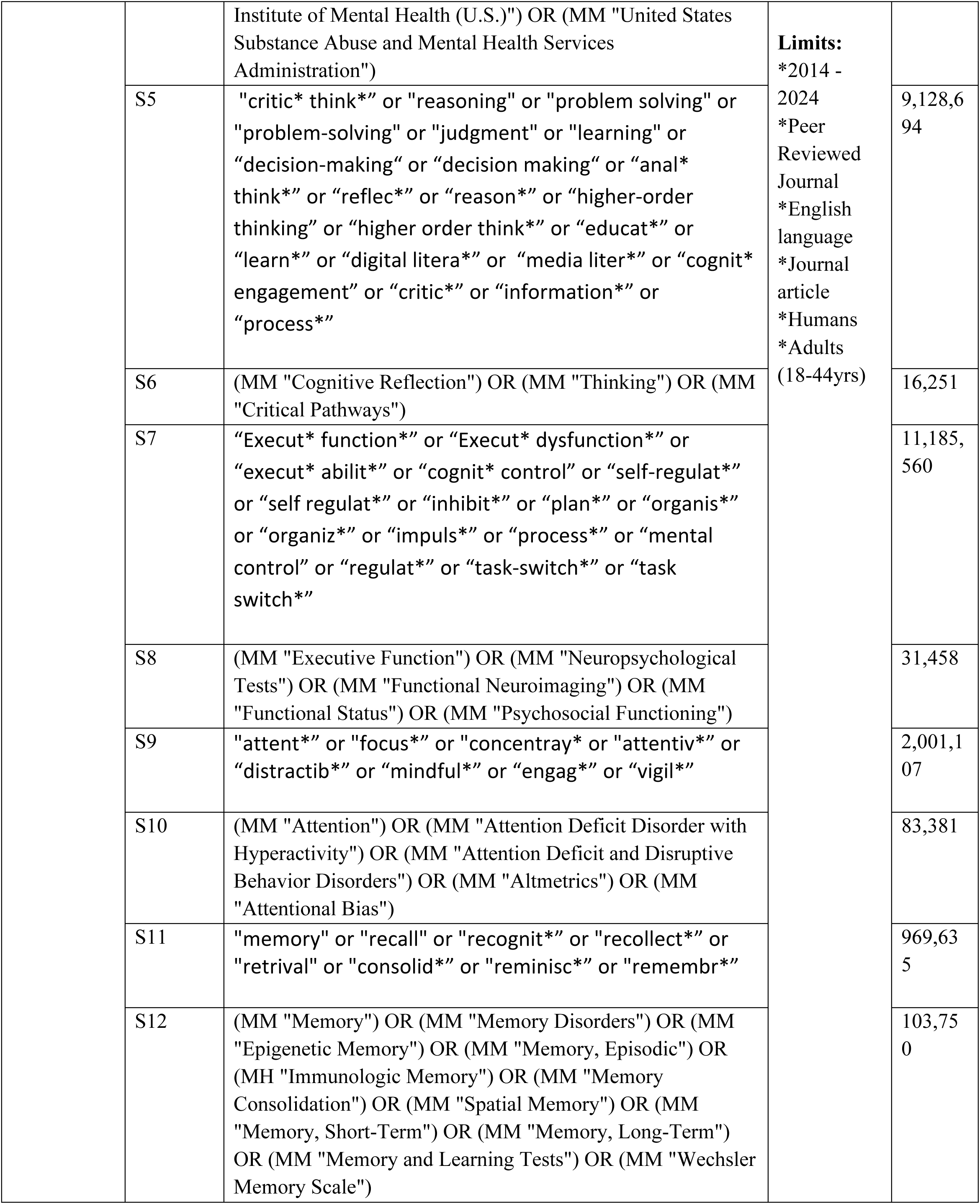
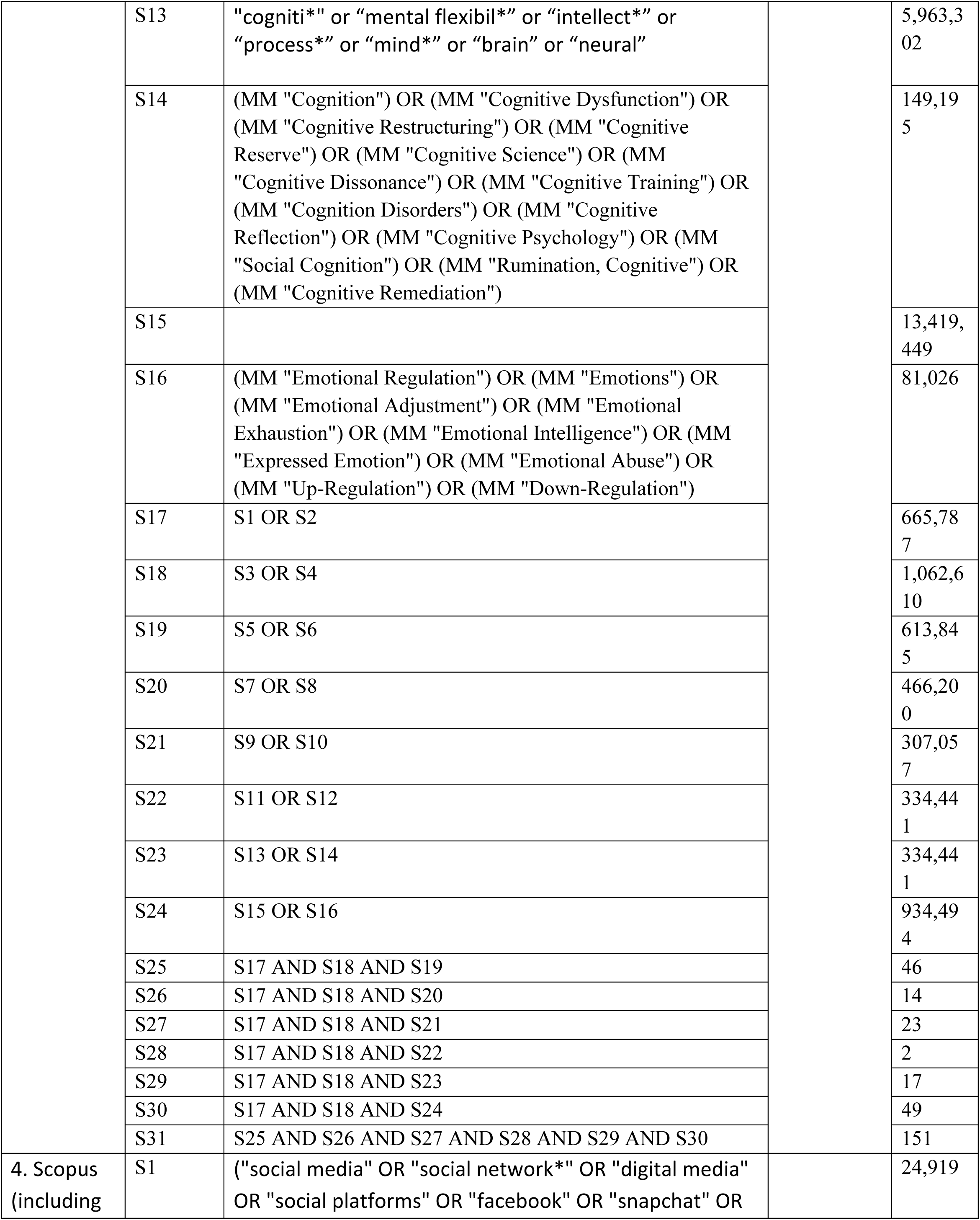
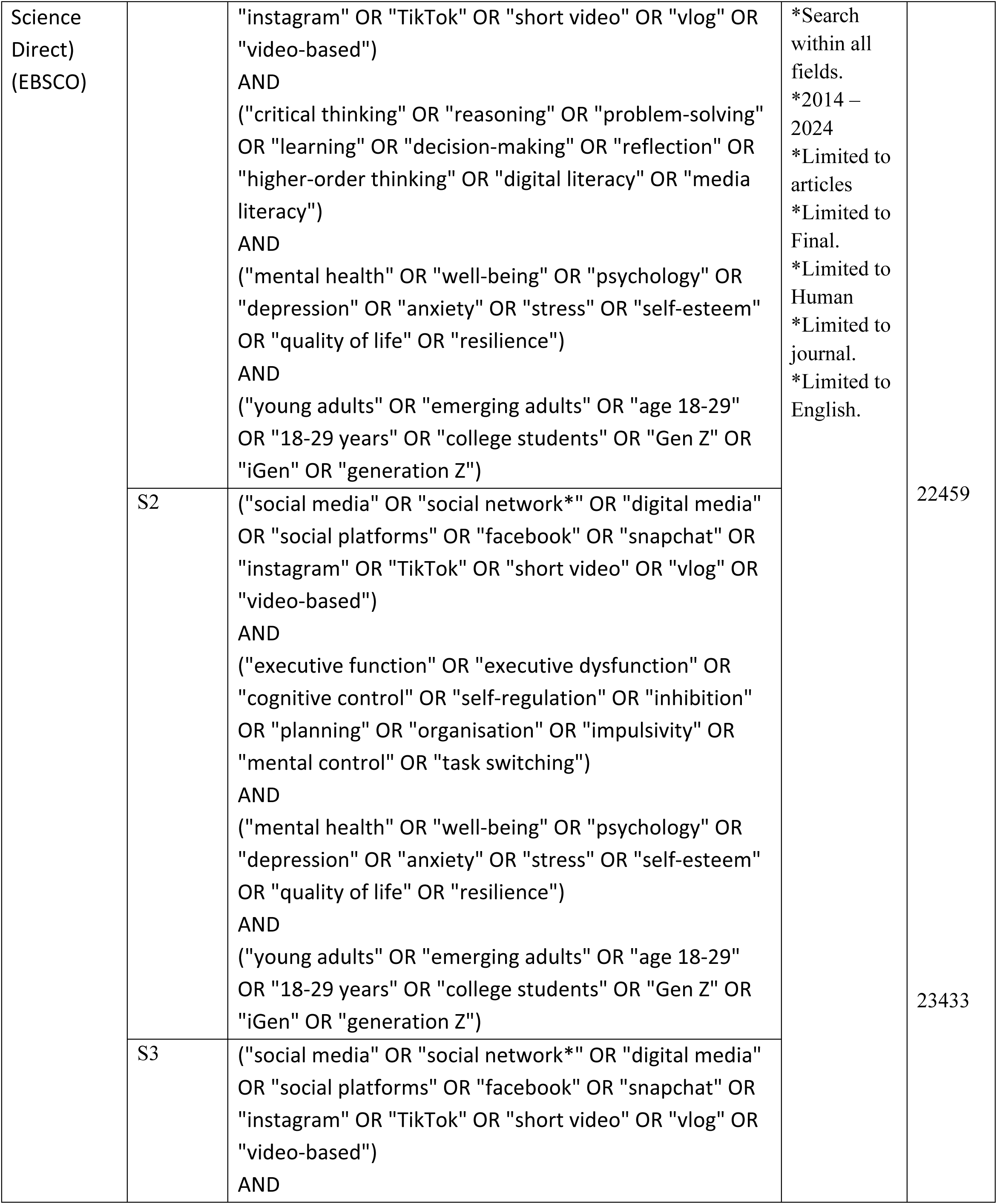
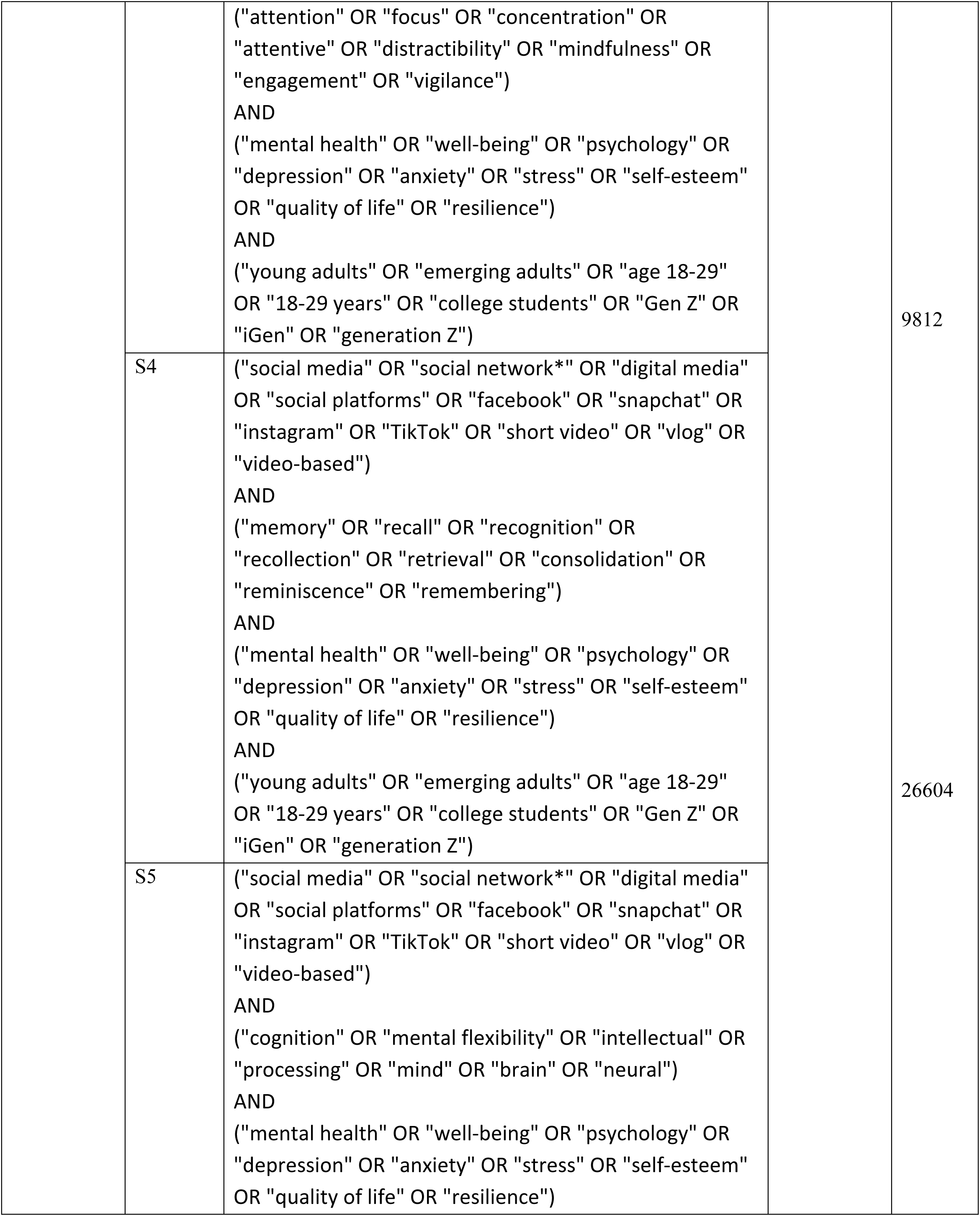
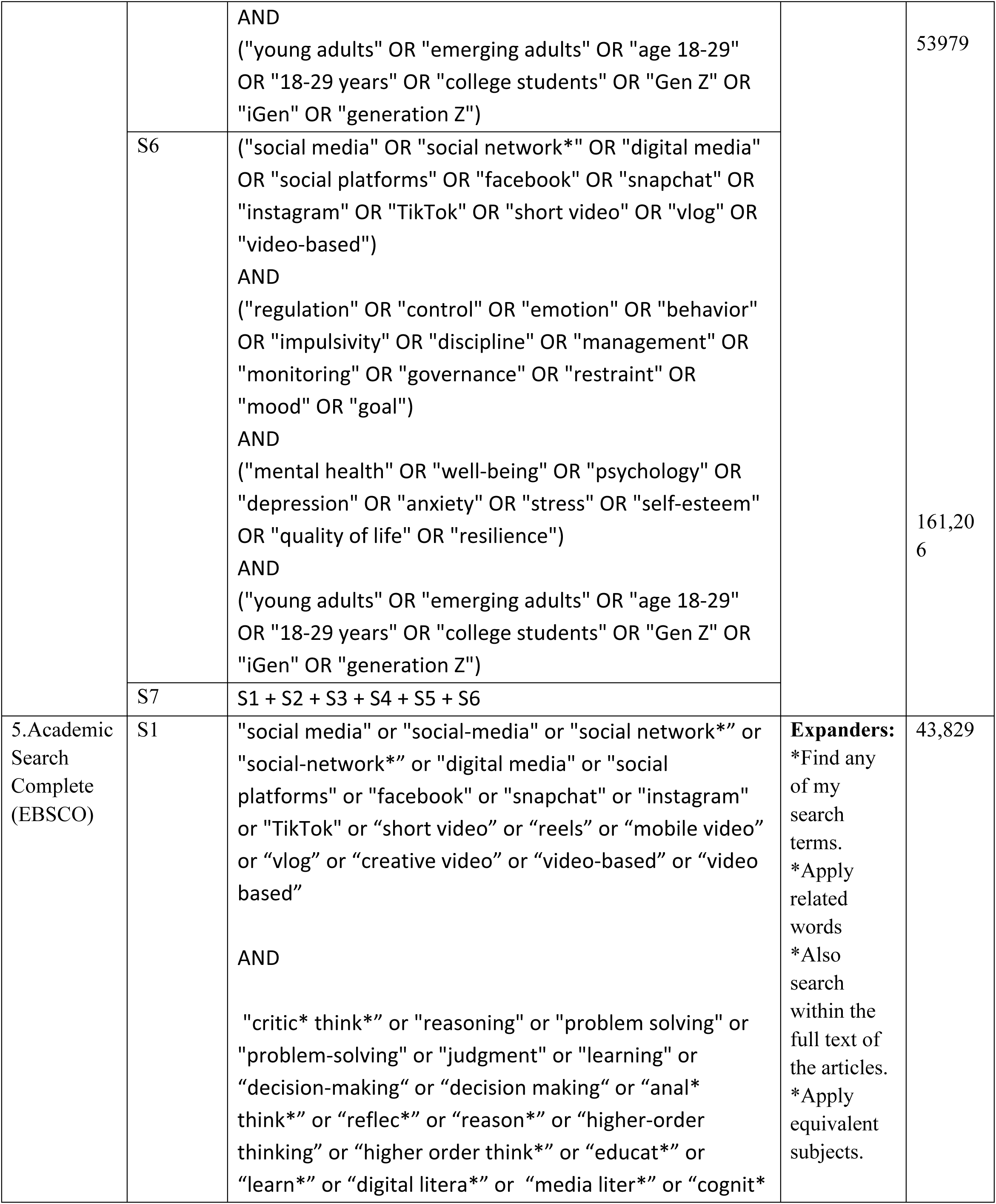
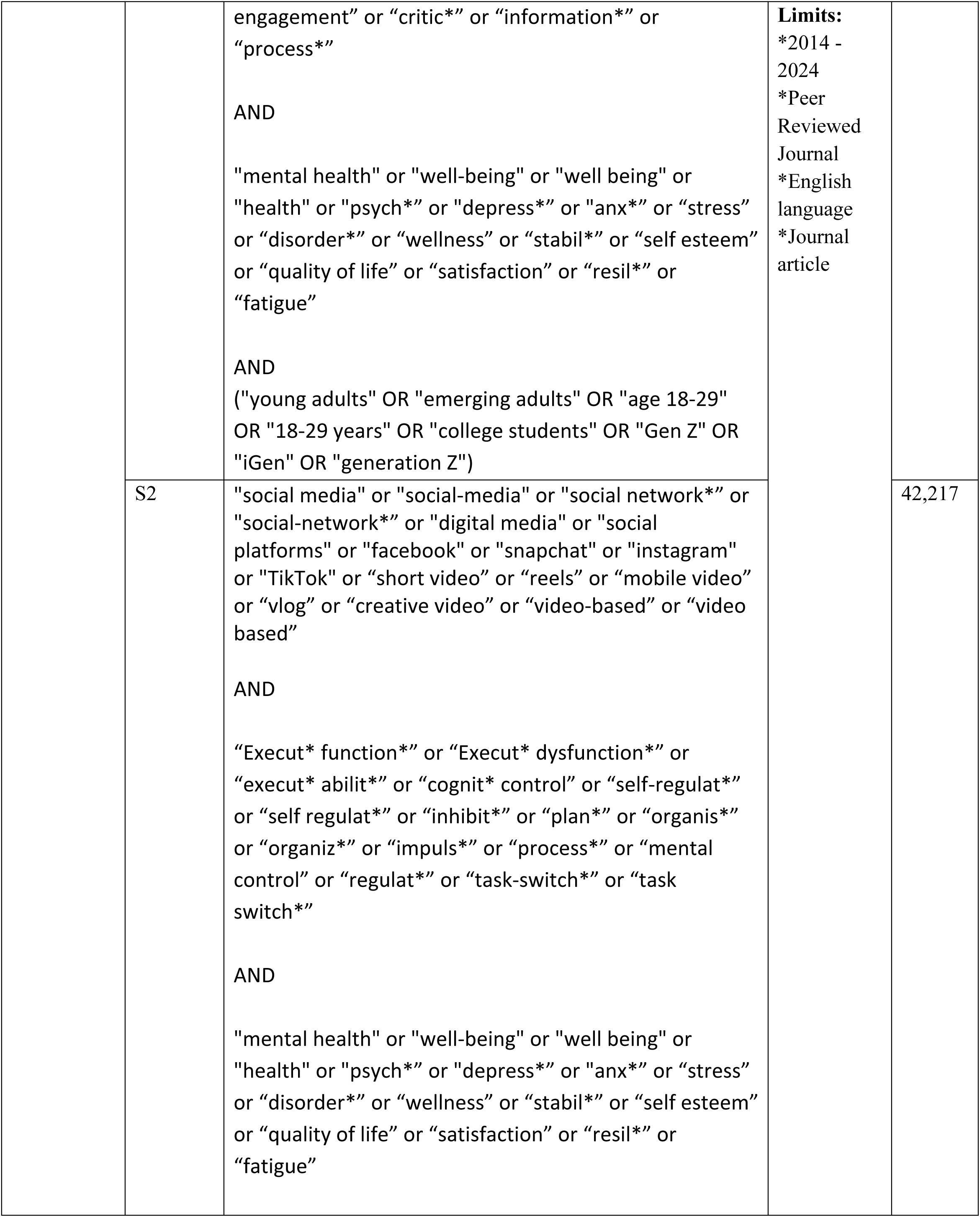
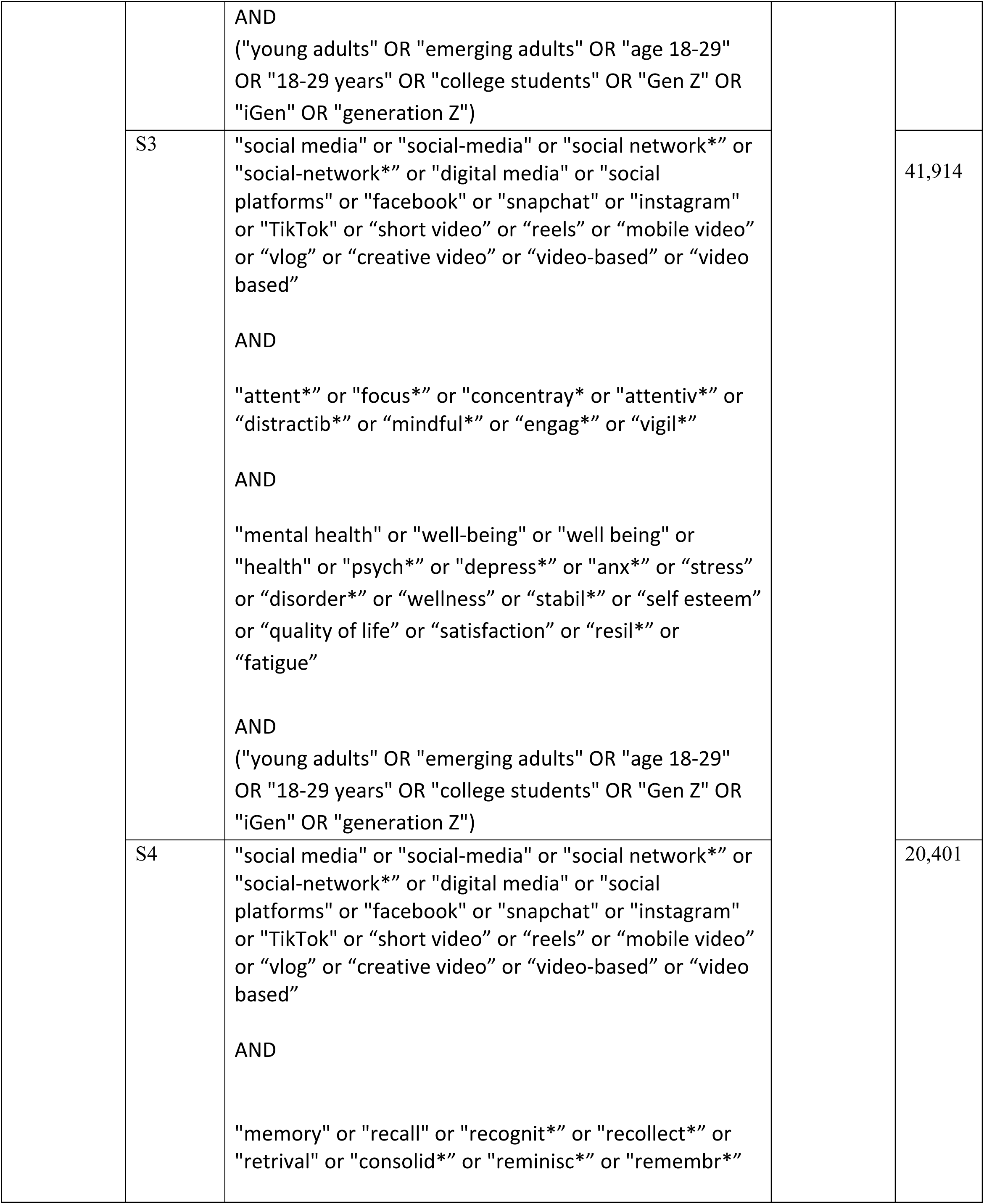
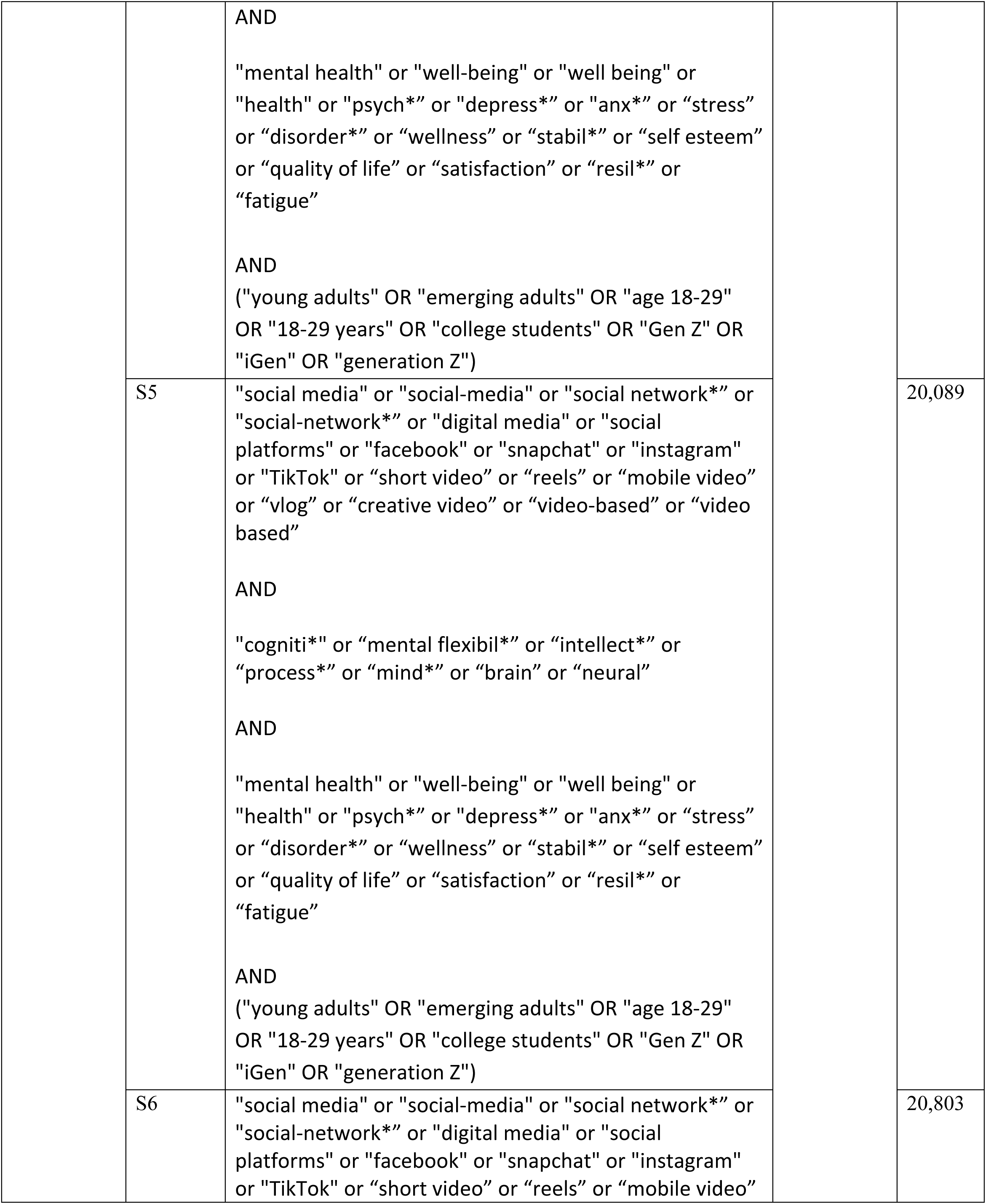
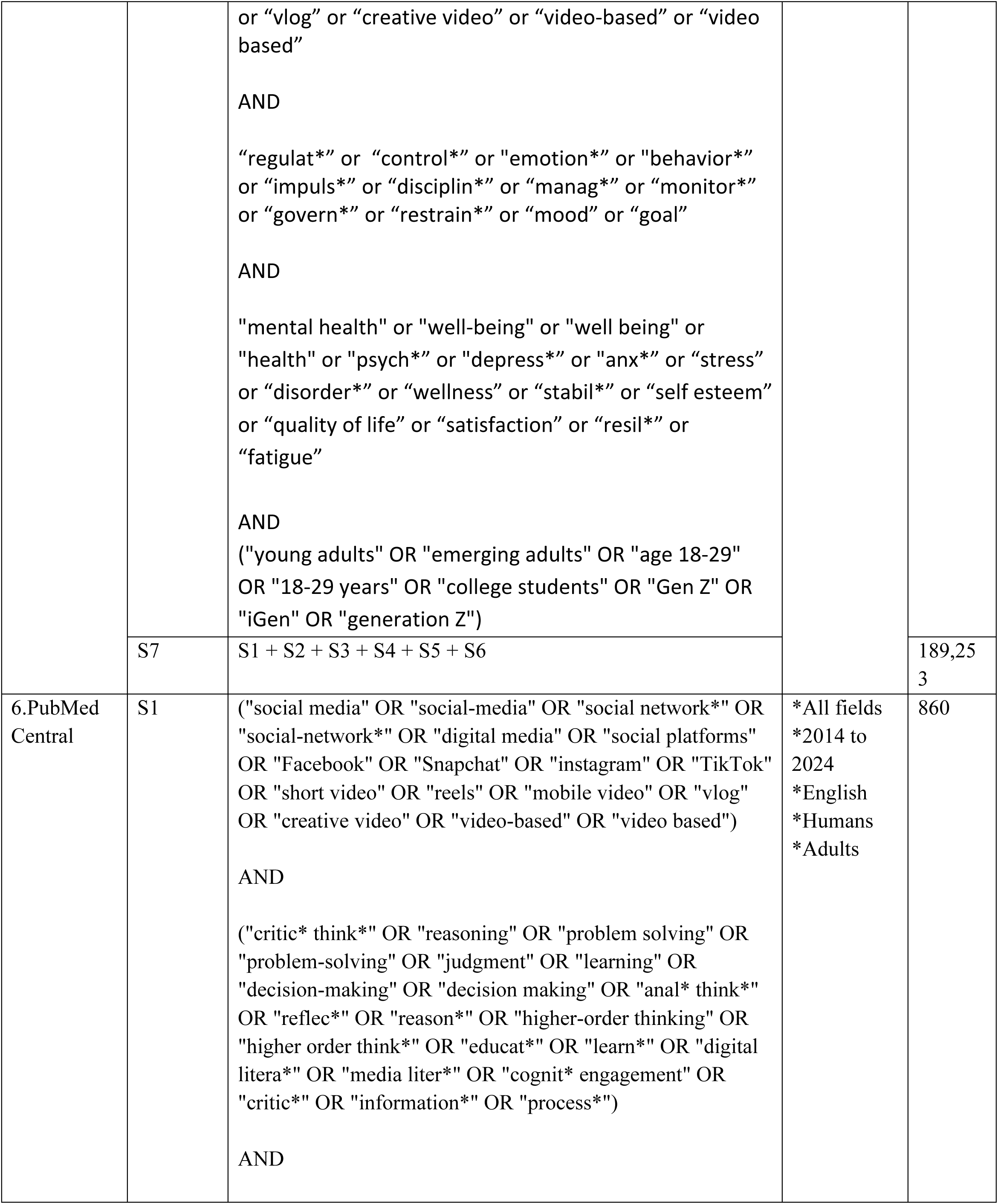
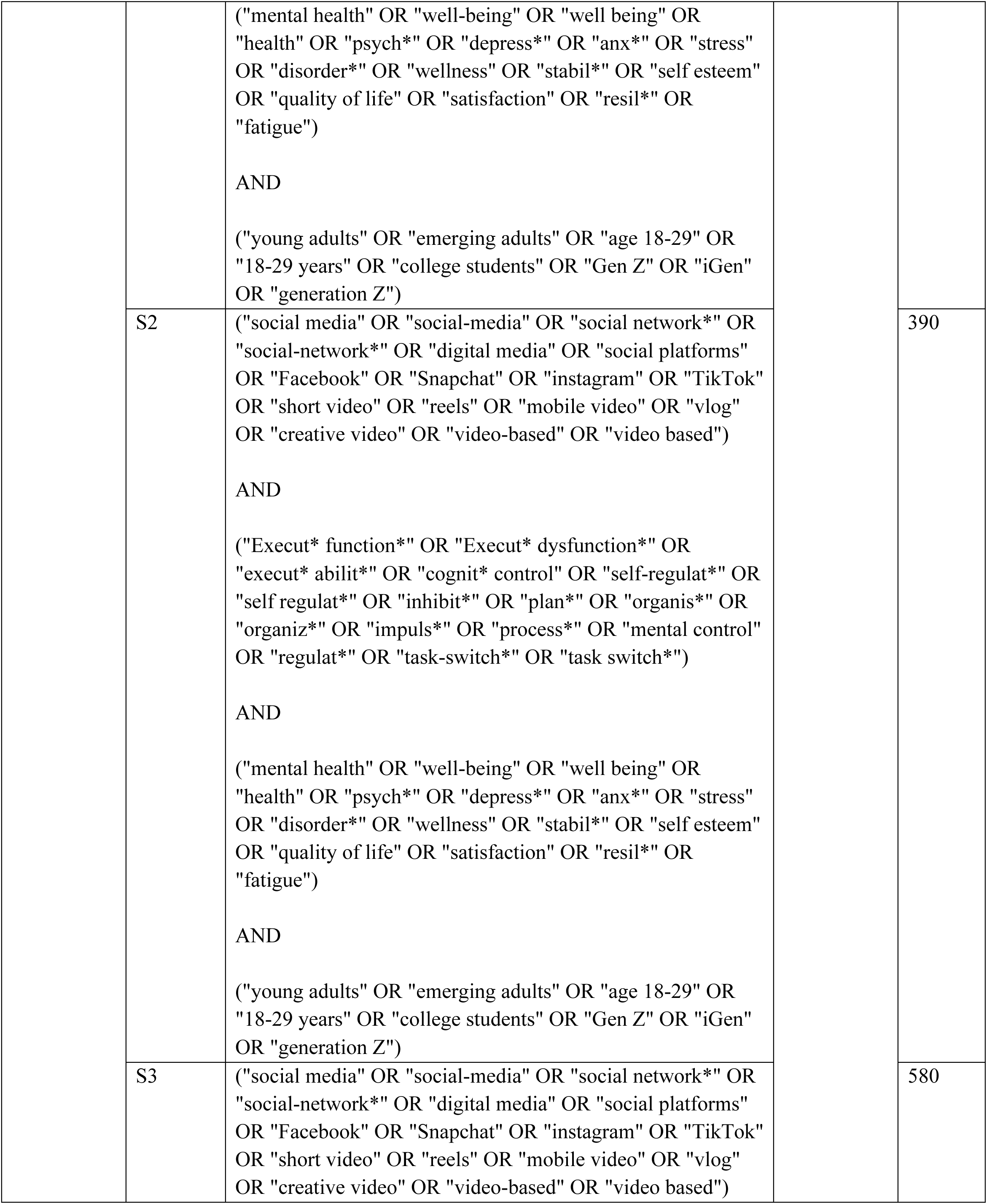
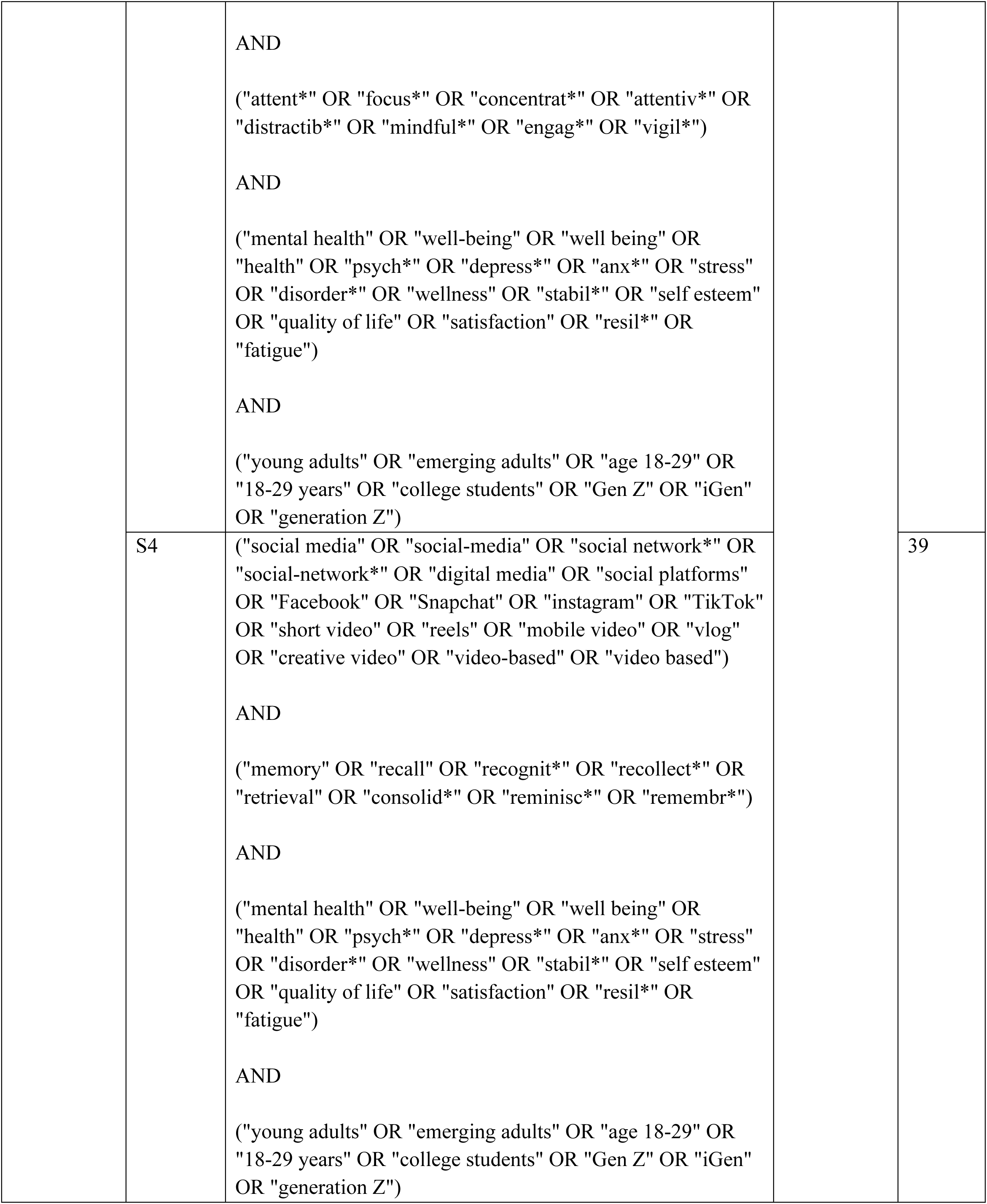
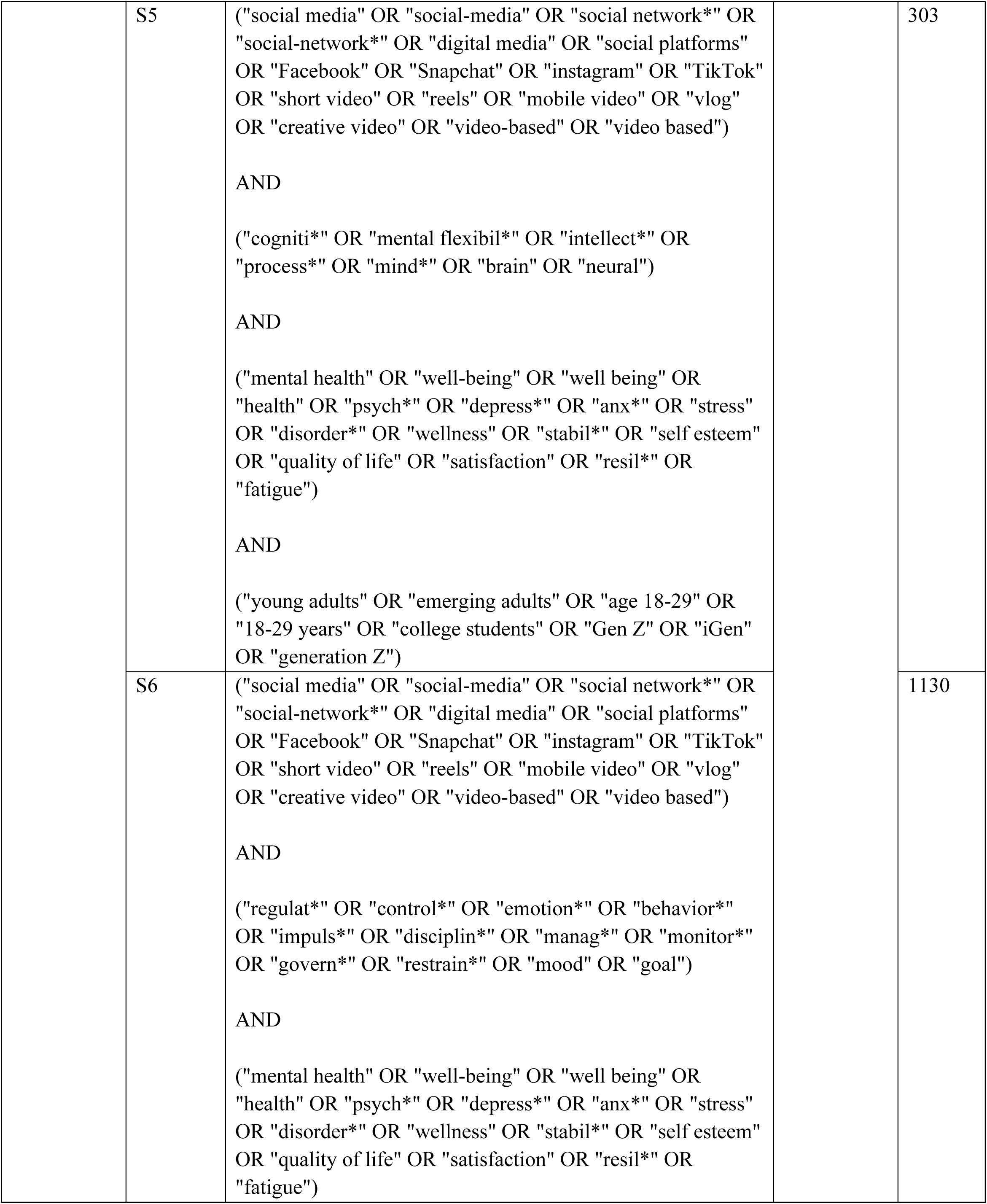
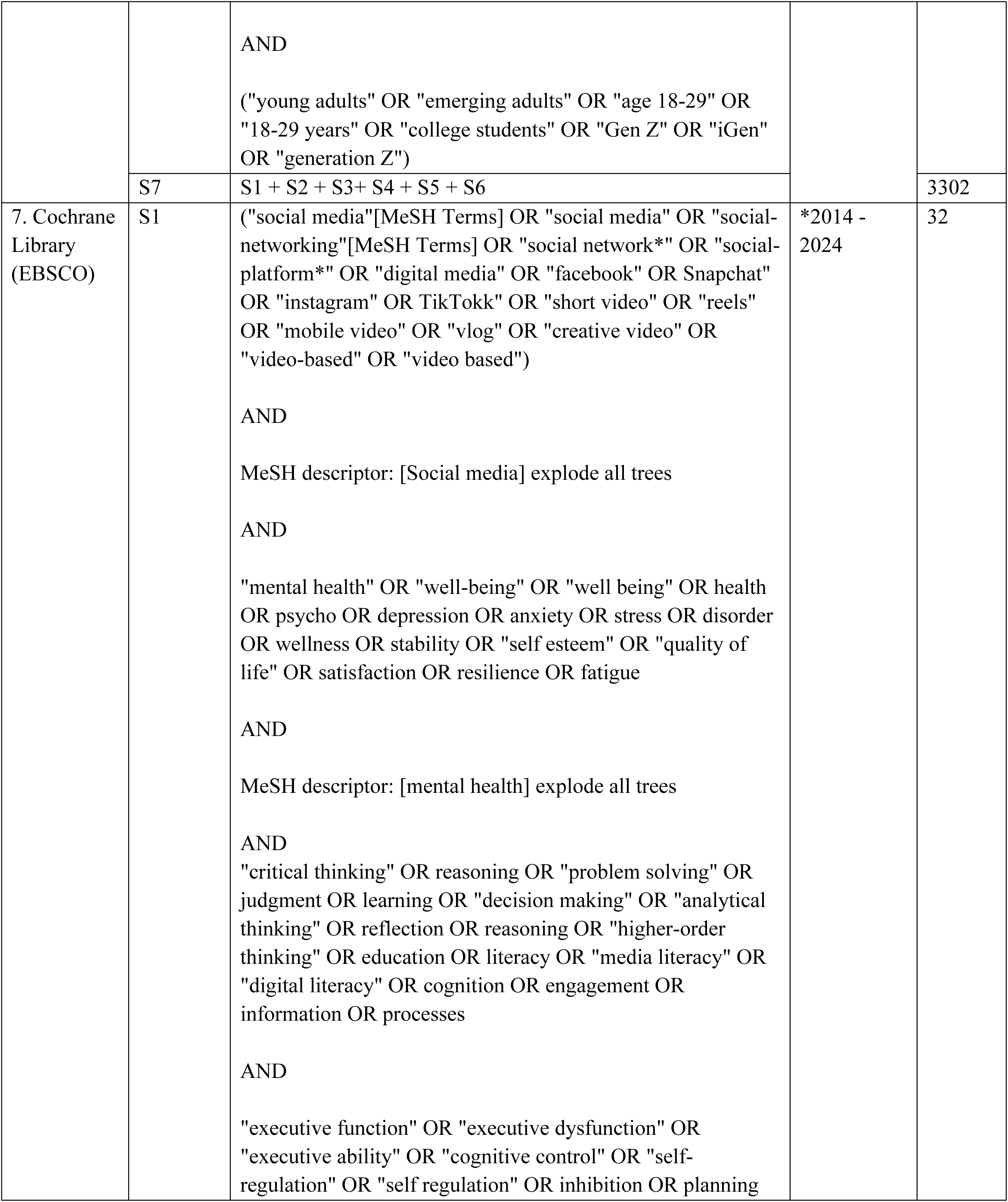
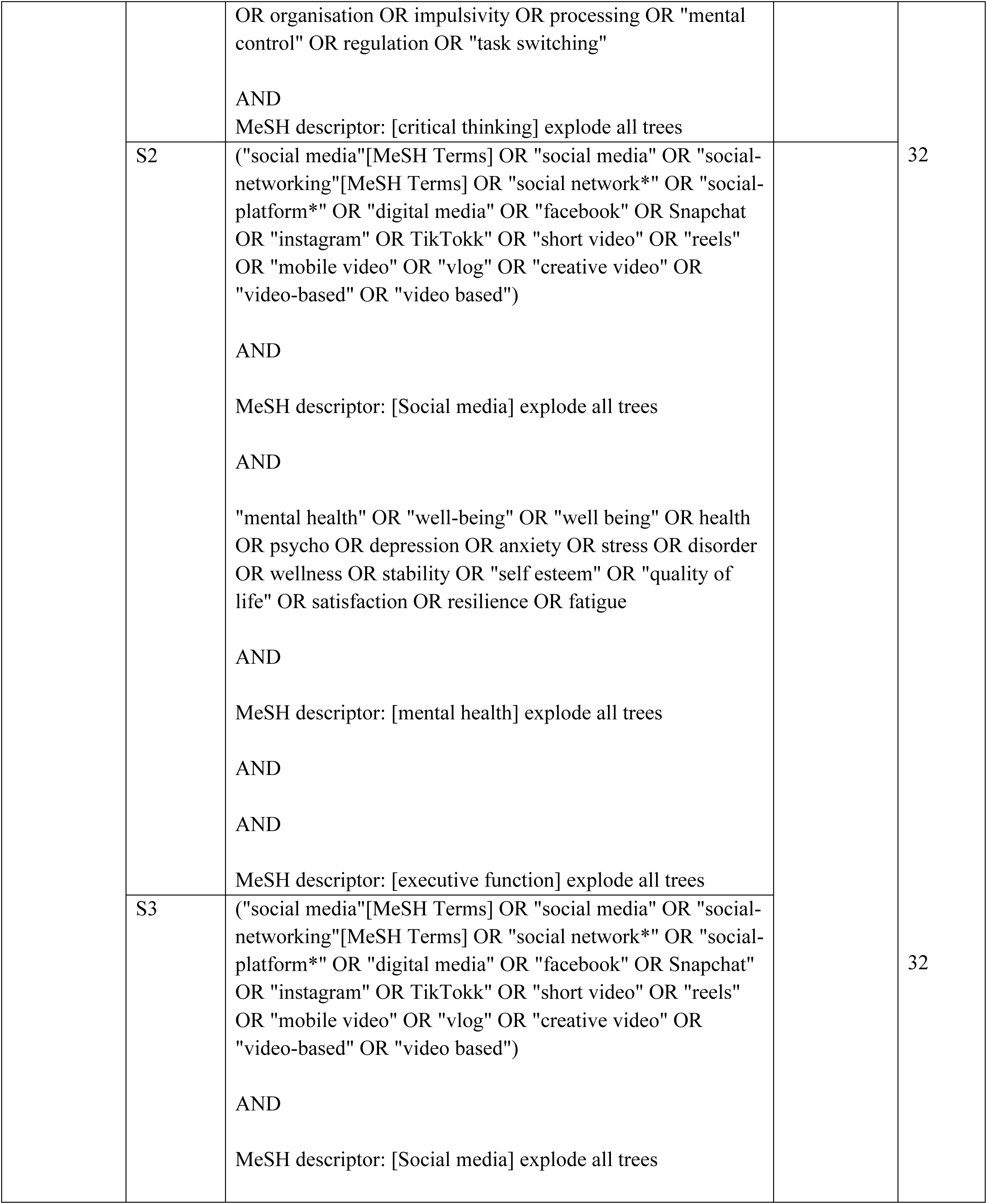
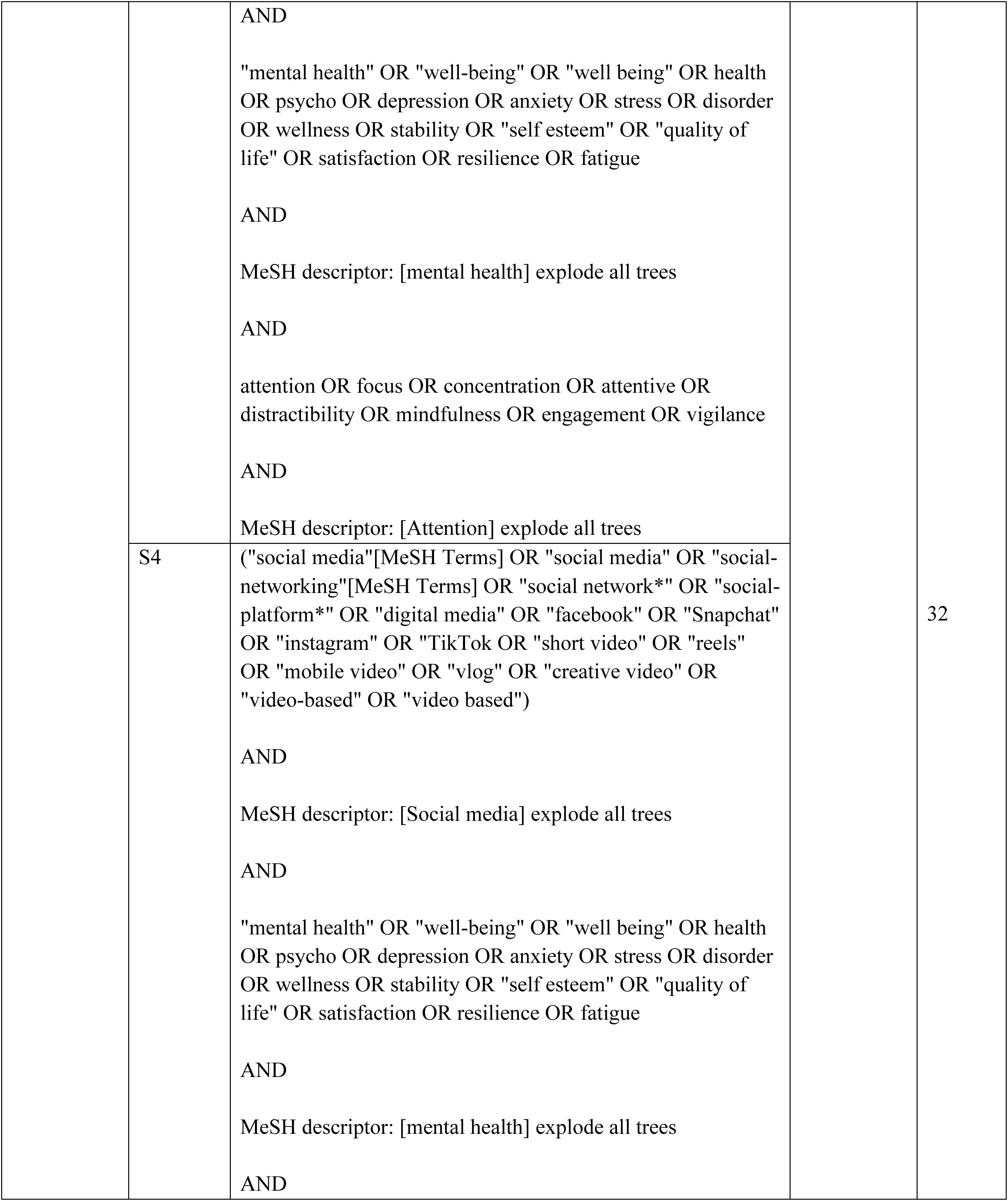
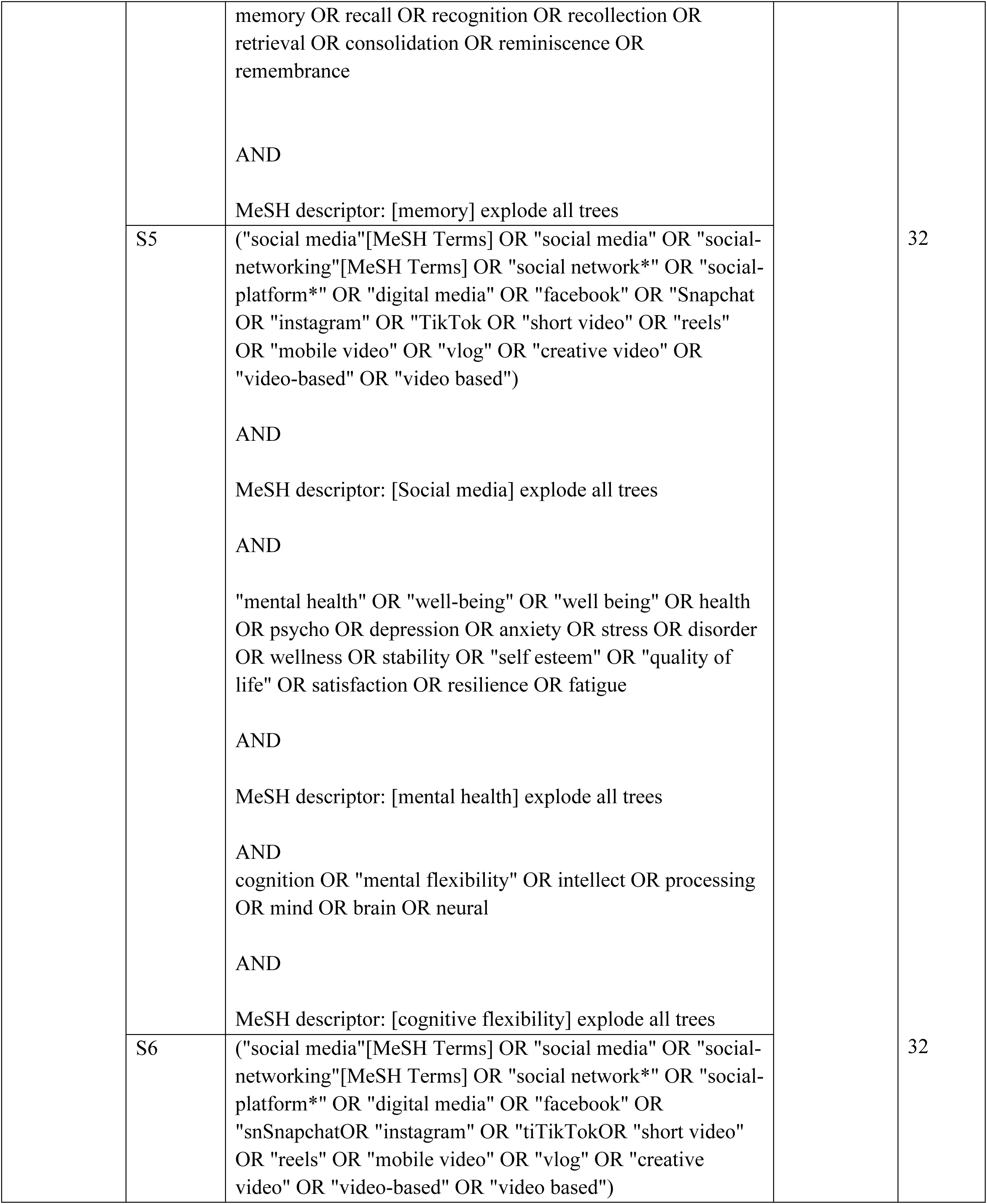
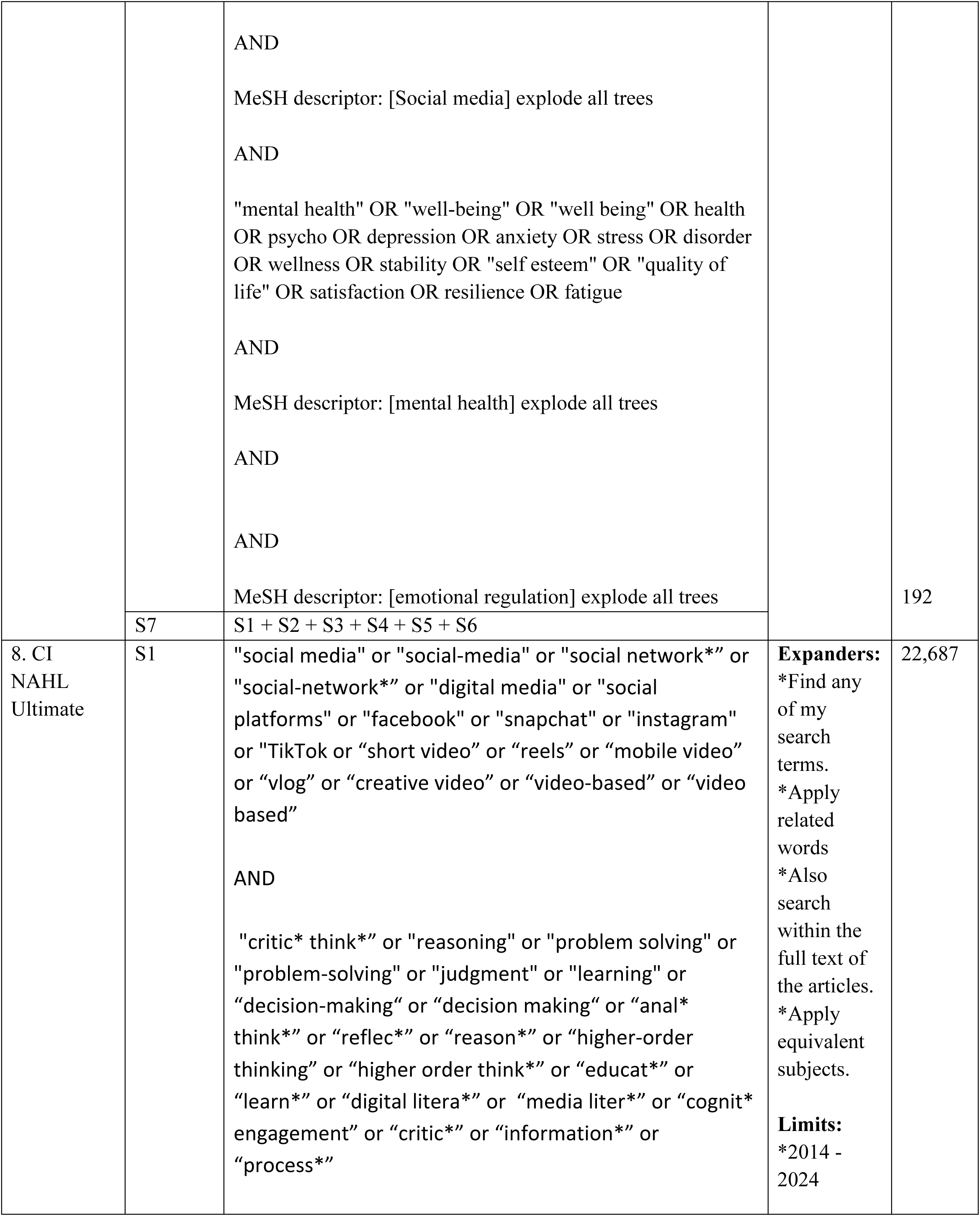
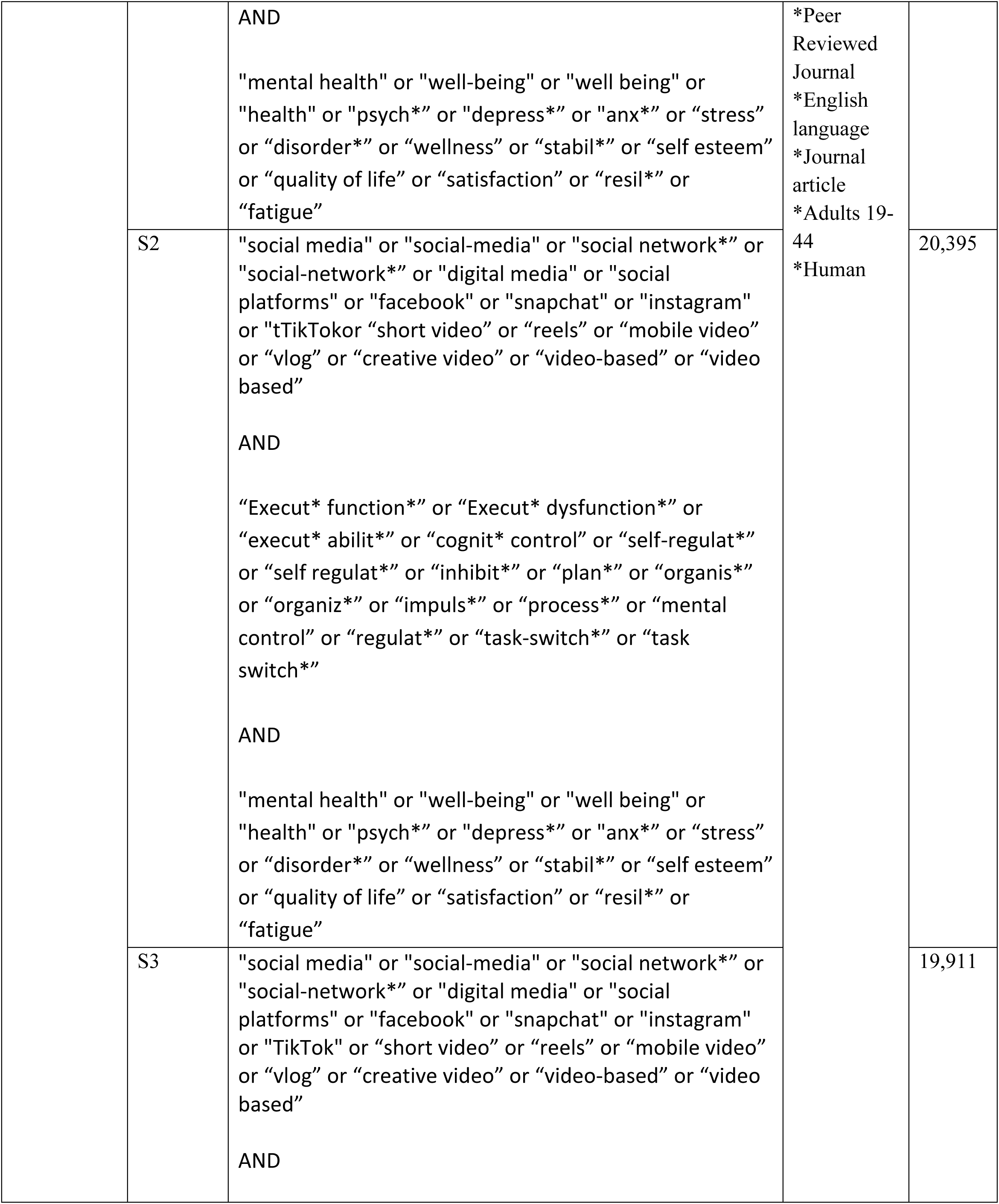
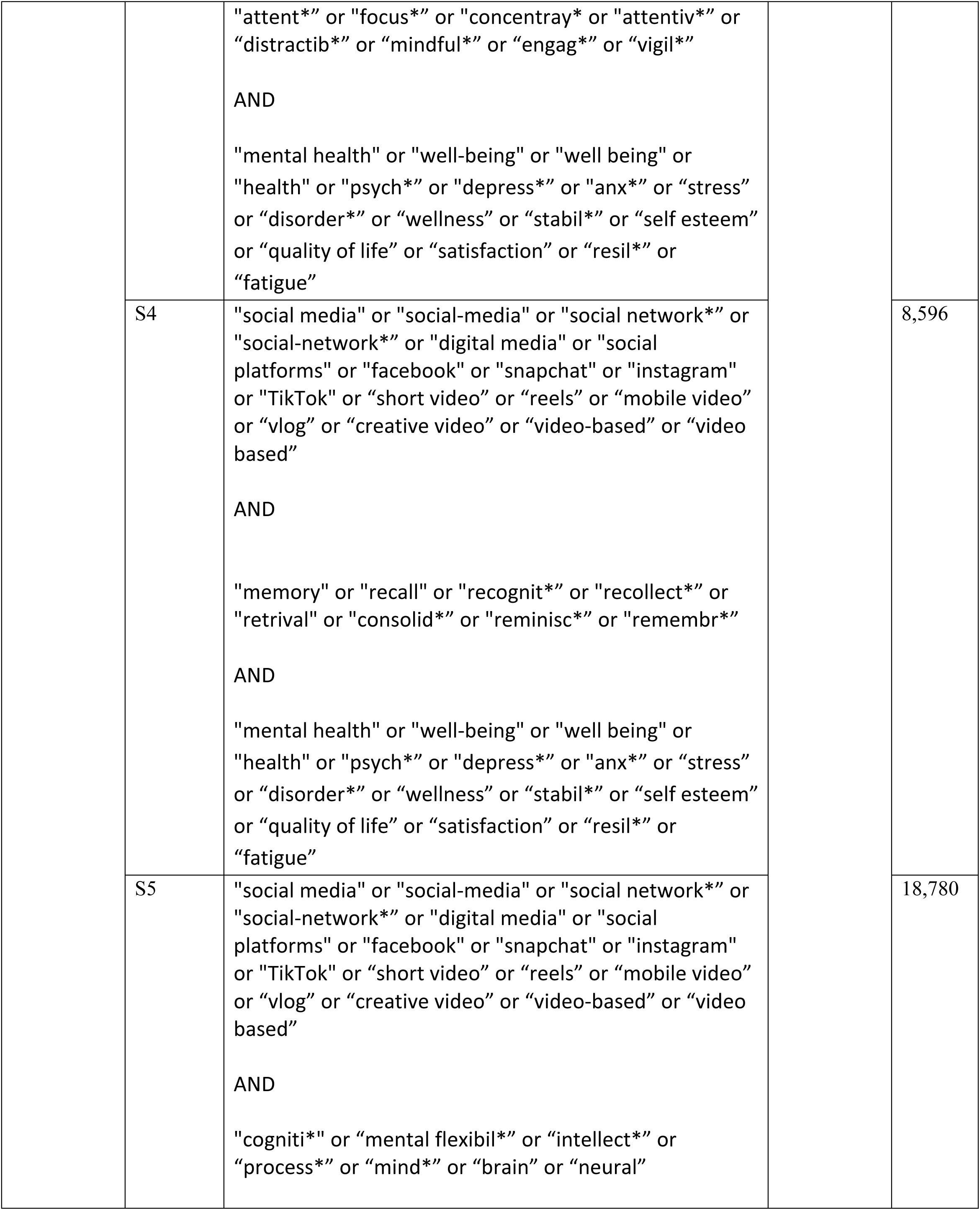
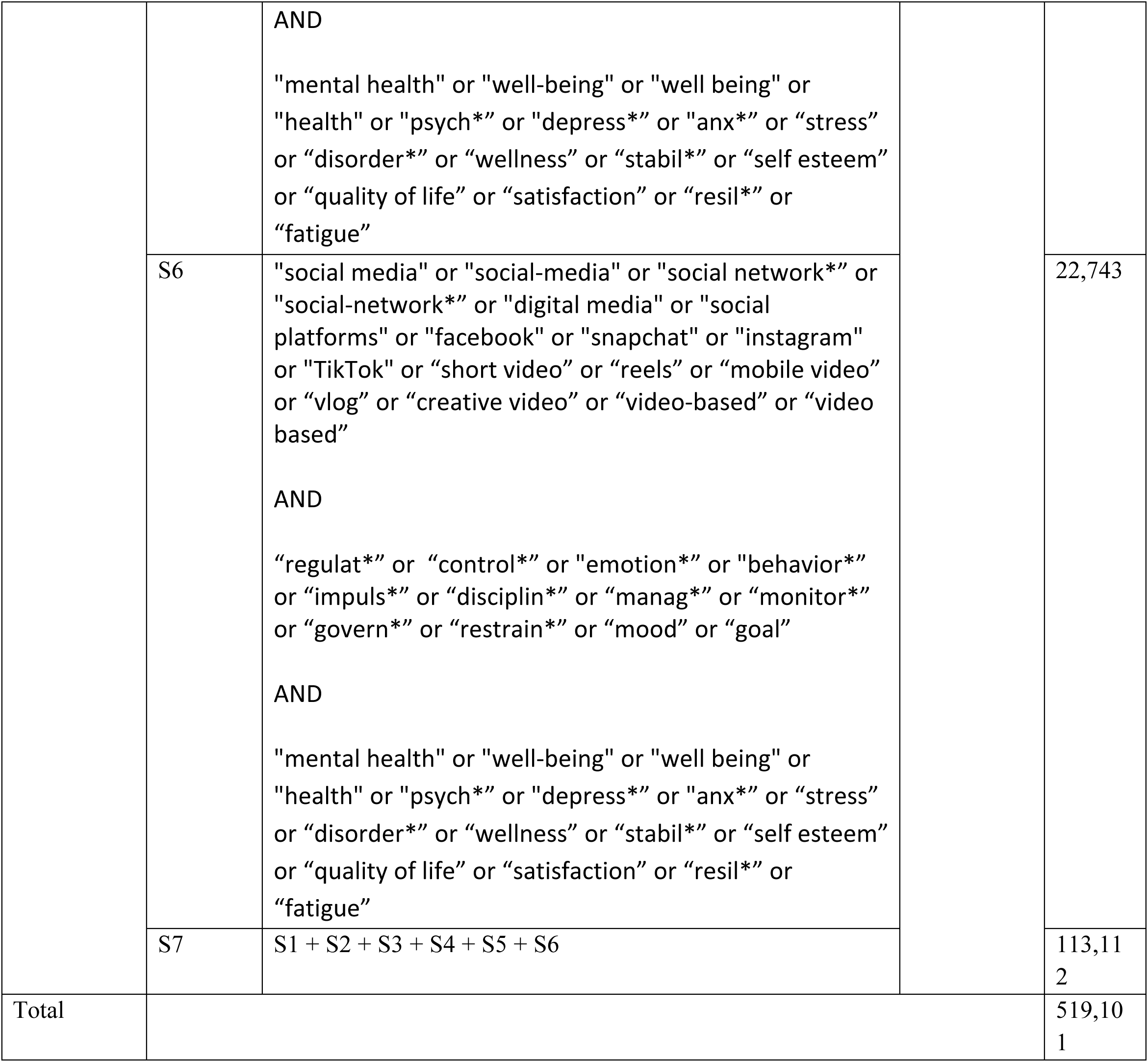
Comprehensive Search Strategy Detailing Keywords, Boolean operators, MeSH words and Databases.

### 2.3. Study Selection

#### Title and Abstract Screening

An initial comprehensive search yielded 519,101 studies. After removing duplicates and applying automated filters based on database restrictions, publication date, peer-review status, keyword relevance, and subject area mismatches, the pool was reduced to 77,678 records. These records were then subjected to title and abstract screening to assess their alignment with our research focus on cognitive mechanisms, mental health impacts, and SFV consumption. During this stage, studies were prioritised according to their methodologies, including the type of SM analysed, the proportion of videos examined, and the measures employed (e.g., scales, questionnaires, interviews). This process refined the selection to 565 studies for full-text review.

#### Full-Text Screening

The full-text screening was completed on the 565 studies based on predefined inclusion and exclusion criteria. This in-depth review ensured that each study adhered strictly to the research focus. Twenty-one studies were identified during this screening process as candidates for further discussion between collaborators. Comments and annotations were added in Rayyan to facilitate a collaborative decision-making process. The next phase involved in-depth reading and highlighting of the selected articles.

#### Exclusion Criteria

Throughout both screening stages, studies were excluded if they met any of the following criteria:

- Investigated populations other than the target demographic (e.g., non-Gen Z groups).
- Employed study designs that were not aligned with the research objectives.
- Focused solely on nutrition or substance use.
- Addressed only subjective well-being without any cognitive function analysis.
- Exclusively examined gaming effects.
- Conducted content analysis solely for intervention creation.
- Investigated interventions for causes unrelated to cognitive or mental health outcomes.
- Explored general SM use without a specific emphasis on SFV content.
- Addressed sleep quality unless directly associated with cognitive function.
- Studied internet-related disorders, social network sites, or general mobile phone behaviour without relevance to mental health and cognition.
- Examined the development or validation of scales related to SM addiction without broader cognitive or mental health implications.
- Researched the effects of SM on personality without focusing on cognitive or mental health outcomes.
- Investigated cultural, political, or familial factors unless they were directly linked to cognitive or mental health.
- Focused on issues such as bullying, drinking, surgery, body image, or eating disorders unless they were connected to social comparison and self-presentation in the context of SFV consumption.
- Assessed FOMO (Fear of Missing Out) and addiction using only basic psychosocial measures.
- Reported mental health outcomes strictly in Gen X or children without relevance to Gen Z.
- Addressed mental health issues (e.g., depression, anxiety) without linking them to cognitive mechanisms.
- Focused on the effects of taking breaks from SM without addressing cognitive outcomes.

#### Snowballing

To enhance the comprehensiveness of the review, snowballing techniques were employed to identify additional relevant studies. This process involved examining the reference lists of the initial 10 included articles, utilising the Research Rabbit tool to explore related literature, analysing Google Scholar citations within referenced works, and reviewing author profiles on the ResearchGate platform. Authors of all included studies were contacted to inquire about any unpublished or overlooked research. After a one-month waiting period, it was found that two of the suggested studies already met the inclusion criteria and had been incorporated into the final selection. One additional study was retrieved through screening of the reference list. In contrast, six further studies were identified through Google Scholar mentions, three of which were initially preprints but had since undergone peer review and publication. All snowballing activities were conducted in conjunction with team discussions to ensure a collaborative and systematic approach to study identification and validation.

**Figure 2.**
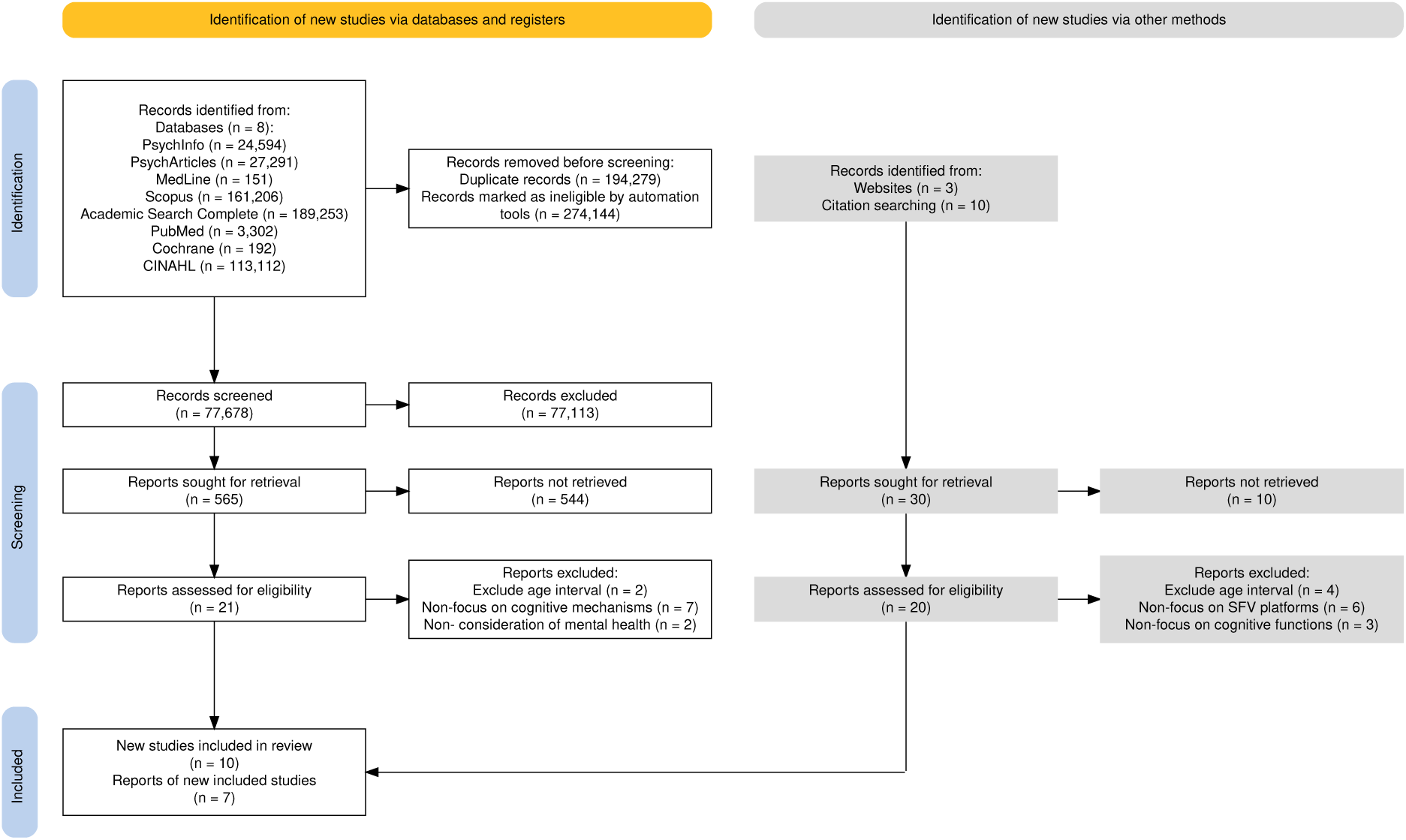
Prisma Flow Diagram illustrating the selection process.

### 2.4. Data Extraction

A full extraction table, including study-level data on participants, interventions, and outcomes, is available in **Supplementary File 1**.

**Figure 3.**
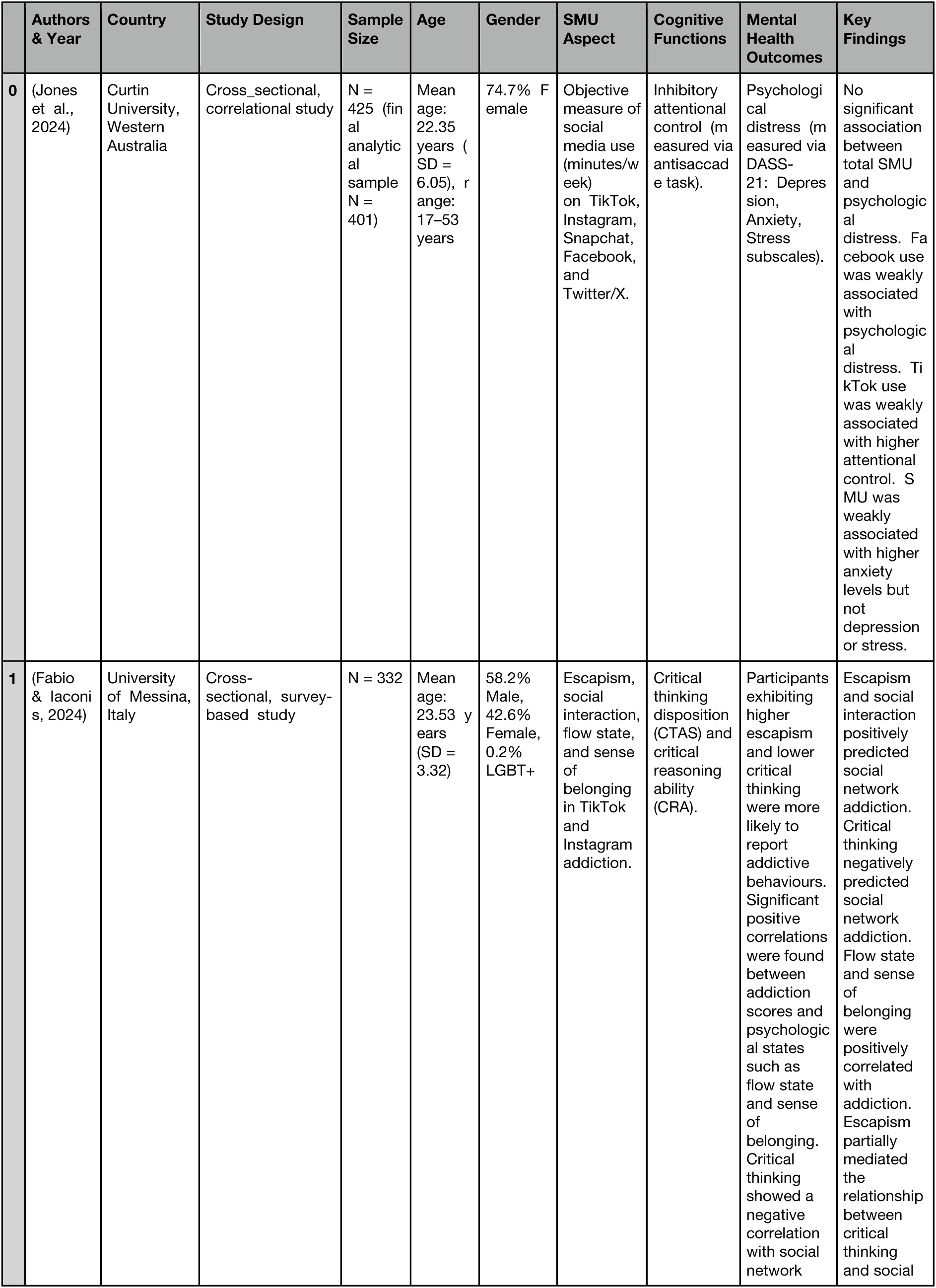
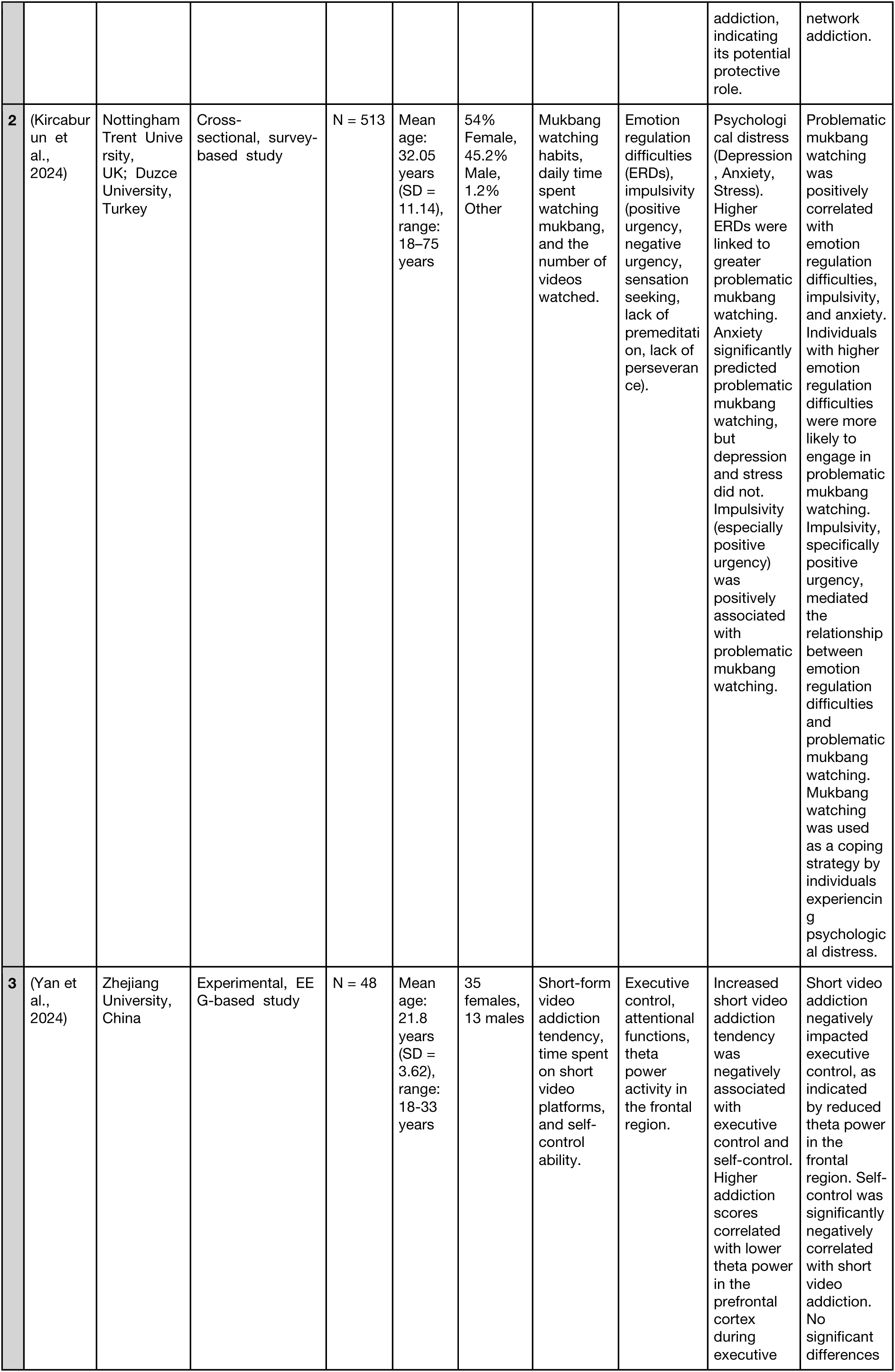
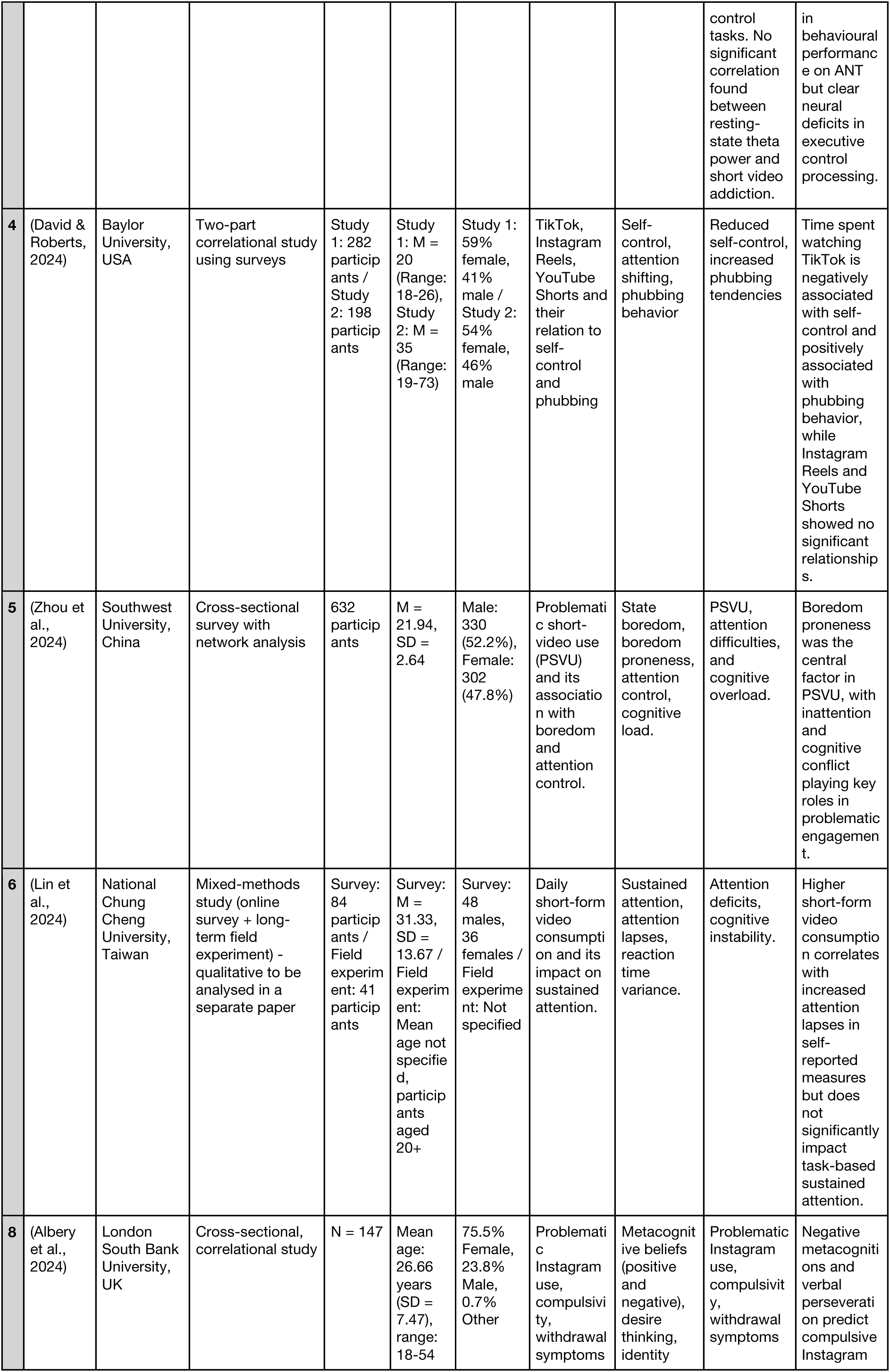
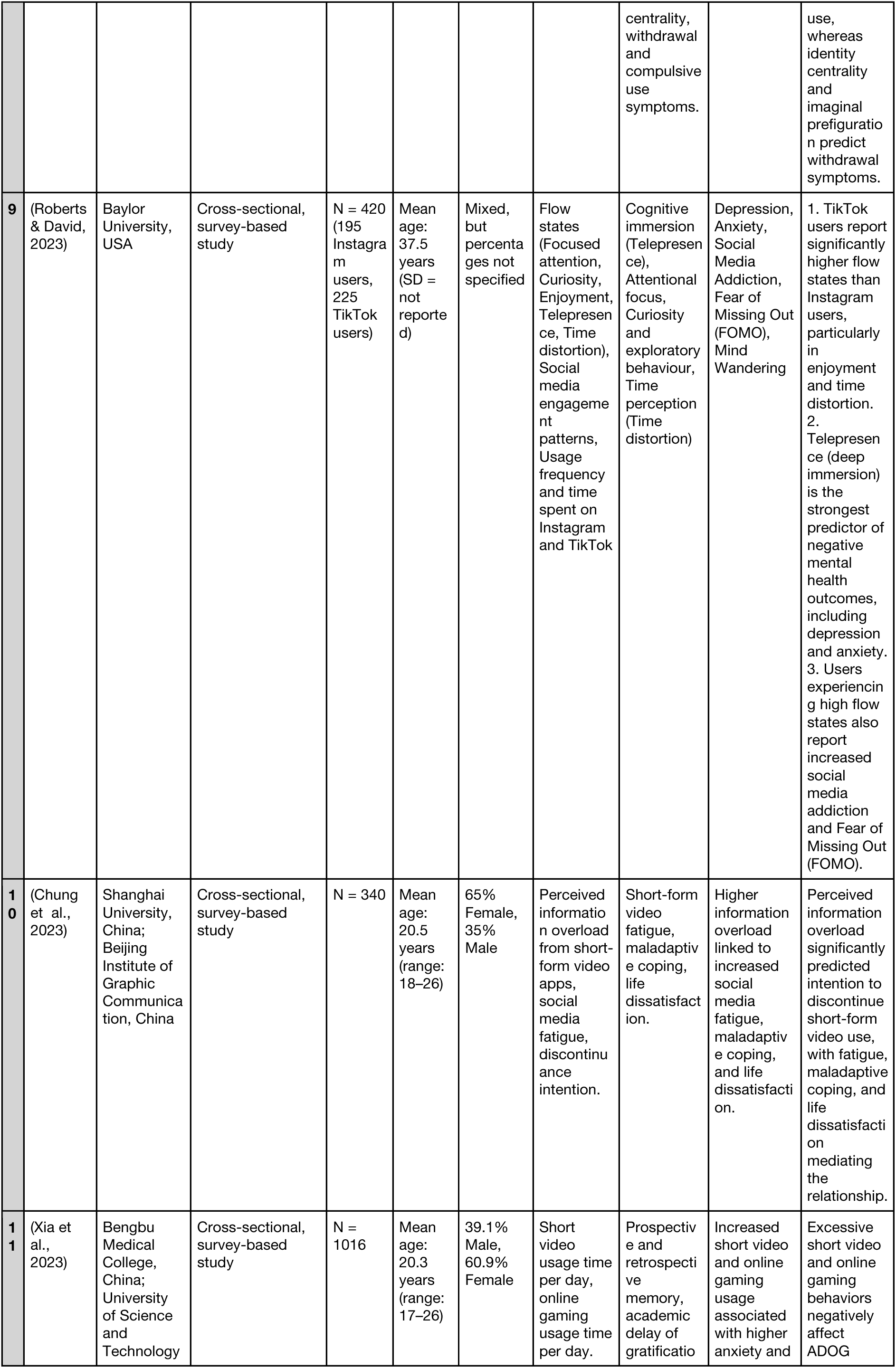
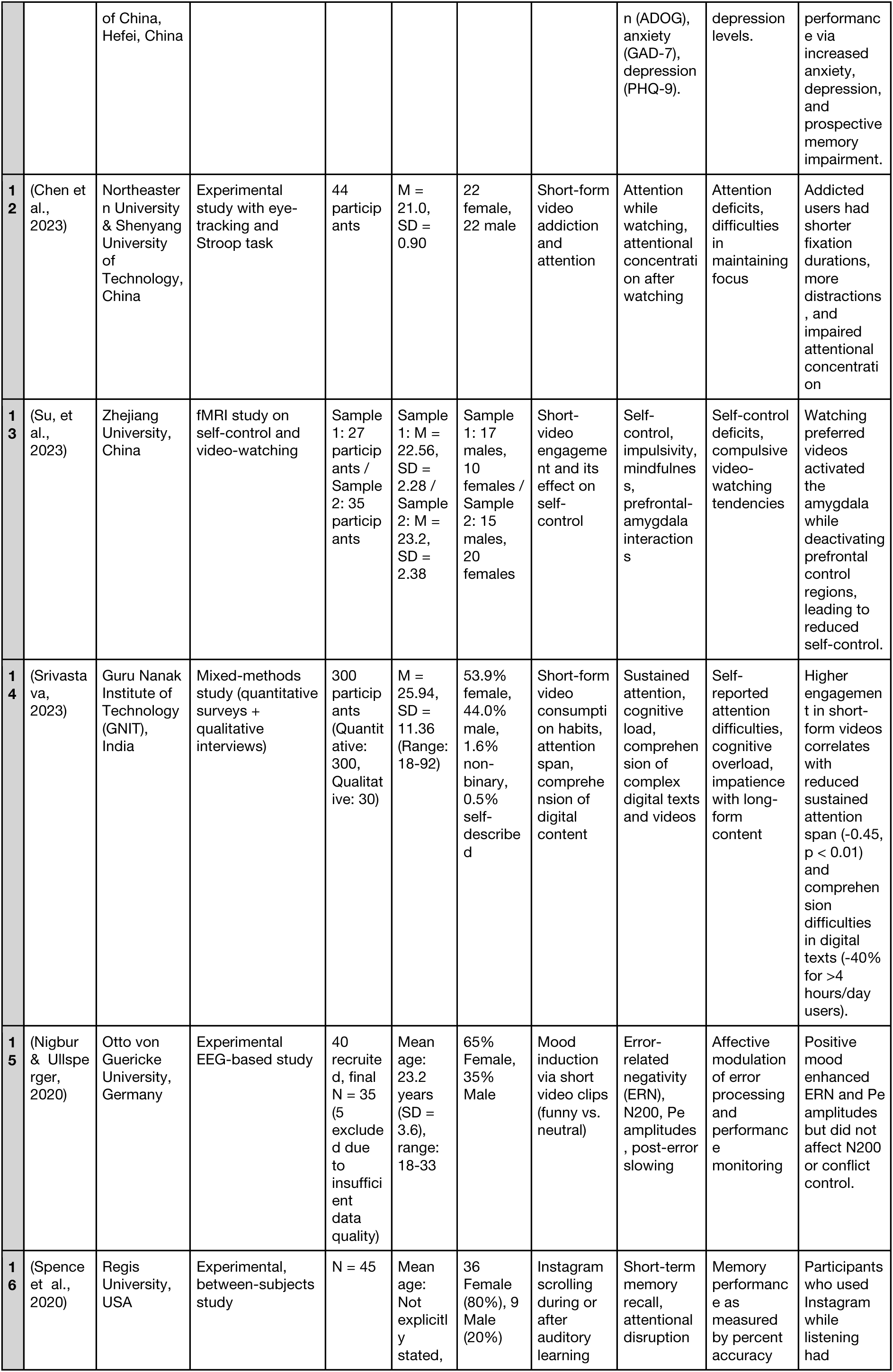
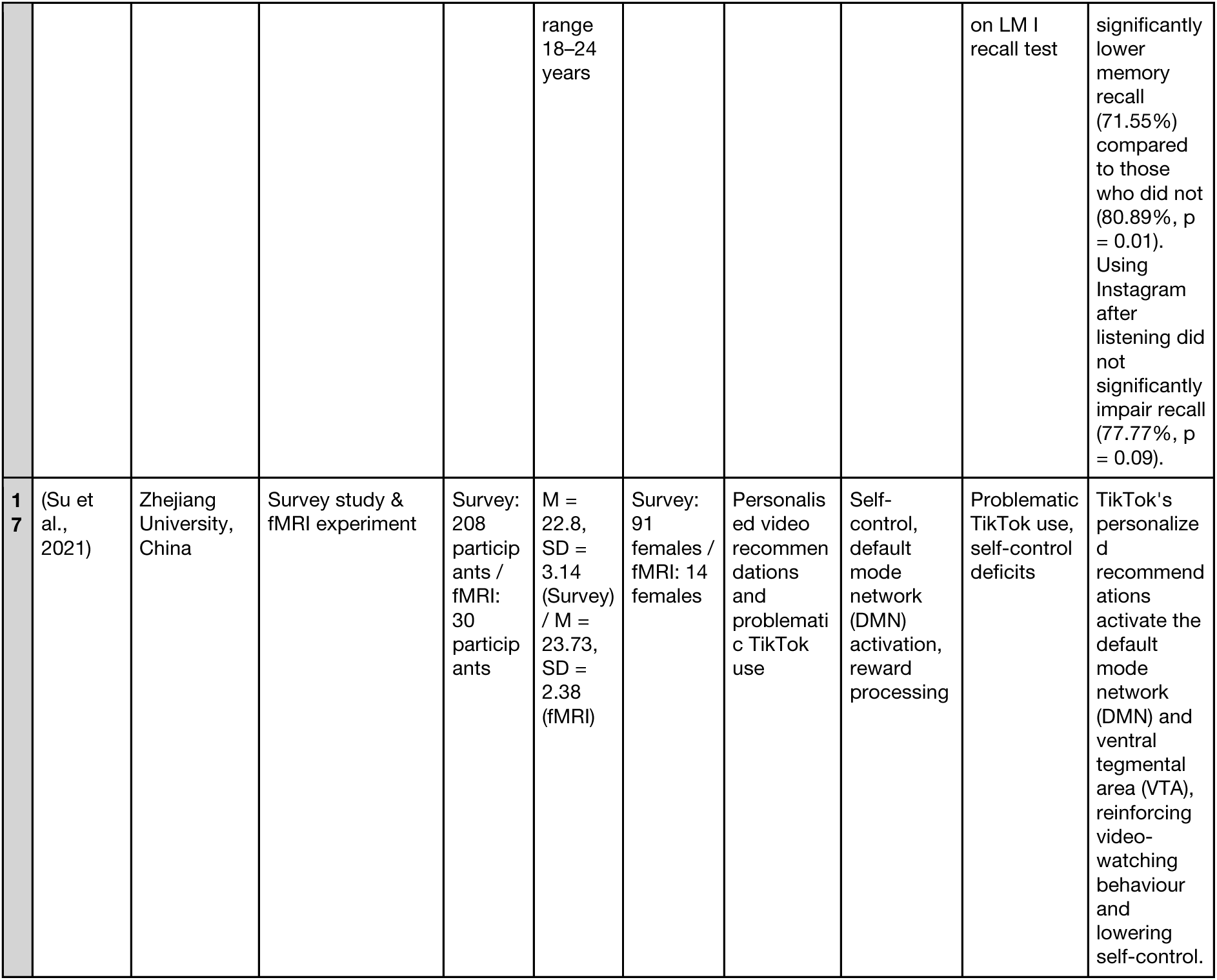
Summary of Data Extraction Table of Included Studies.

### 2.5. Risk of Bias Assessment

Methodological quality was assessed using the Joanna Briggs Institute (JBI) Critical Appraisal Checklist for Analytical Cross-Sectional Studies (Moola et al., 2020). Studies were scored out of 10 based on criteria such as sampling, measurement validity, control of confounders, and statistical analysis. These scores supported the synthesis by highlighting the reliability and limitations of the included evidence.

## 3. Data Synthesis

Building on the outcomes of the systematic search and the structured data extraction process, this synthesis critically examines the emerging literature on SFV use, focusing on cognitive mechanisms and mental health outcomes among Gen Z.

Existing studies point to a paradoxical relationship between SFV engagement, cognitive mechanisms, and mental health. On the one hand, SFV content may enhance mood through short-term emotional stimulation (Nigbur et al., 2024). Conversely, its overstimulating and fragmented format is associated with attentional depletion, diminished impulse control, and maladaptive behaviours such as digital escapism and phubbing (David & Roberts, 2024; Zhou et al., 2024). These concerns are particularly relevant for Gen Z, whose developmental context is shaped by immersive digital exposure.

The present synthesis is organised thematically and methodologically to map how SFV platforms intersect with key cognitive mechanisms explored in the literature—executive function, critical thinking, attention, memory, and emotional regulation—and how these may influence or interact with mental health indicators such as depression, anxiety, and general well-being. Although six cognitive mechanisms were initially targeted—executive function, critical thinking, attention, memory, emotional regulation, and cognitive flexibility—only five consistently emerged across the included studies. Cognitive flexibility was excluded from the synthesis due to the lack of sufficient direct evidence in the final dataset. The structure follows a dual framework, first evaluating cognitive mechanisms and then exploring psychological outcomes while highlighting methodological tensions across the reviewed studies. It integrates findings into a coherent narrative, informing future research.

### 3.1. Cognitive Mechanisms

This section examines the potential impact of consuming SFV on the cognitive mechanisms discussed. It integrates findings from both experimental and self-report studies, comparing their methodologies and outcomes to determine whether SFV usage fosters cognitive engagement or contributes to cognitive overload and mental fatigue.

#### 3.1.1. Executive Functions

A growing body of research suggests that SFV use may compromise executive functioning. By combining neurophysiological, behavioural, and self-report data, studies reveal impaired cognitive control patterns across brain activity and daily experience.

##### Experimental and Neurophysiological Studies

A subset of studies employed experimental paradigms and neurophysiological methods to investigate the impact of SFV use on executive control, the cognitive processes involved in inhibition and conflict resolution. These studies employed objective tools, including EEG, fMRI, eye-tracking, and behavioural tasks, to investigate how SFV may impact executive control and overall functioning.

##### Study (1): Inhibitory Control via Anti-Saccade Task

Researchers employed the anti-saccade task to assess inhibitory attentional control, a fundamental component of executive functioning. Contrary to initial expectations, the study found no negative association between total SM use and inhibitory control. TikTok use was weakly positively associated with better inhibitory control accuracy, suggesting that frequent engagement with SFV content may not impair but rather slightly enhance certain aspects of attentional regulation (Jones et al., 2024).

##### Study (4): Executive Attention via EEG-Based Attention Network Task

Another study employed the Attention Network Task (ANT) in conjunction with Electroencephalogram (EEG) to assess three attentional components: alerting, orienting, and executive control. While no behavioural deficits were observed, individuals with higher scores on the SM addiction questionnaire (adapted to SFV) showed reduced theta power in the frontal region during conflict trials. This neural marker suggests compromised executive control, particularly in the brain’s capacity to resolve conflict and sustain attention. These findings suggest a potential neural-level disruption in cognitive regulation associated with excessive SFV use (Yan et al., 2024).

##### Study (17): fMRI and Multimodal Self-Regulation Profiling

Similarly, researchers employed Functional Magnetic Resonance Imaging (fMRI) and self-report scales to assess executive function and self-control. Neuroimaging results revealed reduced activation in the Dorsolateral Prefrontal Cortex (DLPFC) and Anterior Cingulate Cortex (ACC) regions, which are critical for executive control and decision-making when participants viewed personalised TikTok videos. Participants with higher scores relating to problematic TikTok use (adapted from the Internet Addiction Test) further reported greater impulsivity and lower self-control, suggesting a neuro-behavioural link between SFV engagement and impaired cognitive regulation (Su et al., 2021).

##### Study (13): Reward-Based Executive Function via fMRI

In their follow-up study, Su et al. (2021) employed fMRI to investigate how SFV cues influence reward-based decision-making and self-regulation. The results revealed that viewing personalised SFV content triggered increased activation in the Ventral Tegmental Area (VTA)—a core region in the brain’s reward circuitry—while simultaneously showing decreased functional connectivity between Default Mode Network nodes and executive control regions such as the Anterior Cingulate Cortex (ACC) and Precuneus. These findings suggest a dual-process imbalance in which impulsive, reward-seeking neural systems may override reflective, top-down self-control mechanisms in individuals who use SFV more frequently.

##### Study (15): EEG, EMG, and Physiological Stress Responses

This study combined Electroencephalography (EEG), Electromyography (EMG), and skin conductance recordings during a conflict-resolution task to examine the influence of emotionally evocative video content on performance monitoring. Participants exposed to humorous short clips—similar to those in SFV—showed increased emotional arousal and increased Zygomaticus EMG activity. Neurophysiological results revealed enhanced error-related negativity (ERN) and error positivity (Pe) following mood induction, indicating heightened emotional reactivity to performance errors. These findings suggest that exposure to emotionally charged SFV content may enhance the affective salience of cognitive errors while weakening top-down regulation in emotionally loaded contexts (Nigbur & Ullsperger, 2020).

##### Synthesis and Relevance

Collectively, these studies suggest that SFV engagement can compromise executive control across behavioural, neural, and emotional dimensions, although effects may vary depending on the task, modality, and emotional context:

| Study | Conclusion |
| --- | --- |
| Jones et al., 2024 | Neutral/Enhanced inhibitory control. |
| Yan et al., 2024 | Reduced frontal theta during conflict trials. |
| Su et al., 2021 | Boosted reward activity and weakened control. |
| Su et al., 2024 | Heightened emotional and error responses. |
| Nigbur & Ullsperger, 2020 | Reduced Executive Activation (DLPFC, ACC). |

##### Survey and Scale-Based Studies

While experimental studies provide insights into neural and behavioural mechanisms, other research has explored executive functions mainly through self-report tools and psychometric scales. These studies assess users’ perceived ability to manage attention, inhibit impulses, and regulate emotions in daily life, providing ecological and psychological context.

##### Study 3: Emotion Regulation and Impulsivity via DERS

Researchers found that problematic mukbang watching can act as a coping mechanism for emotional distress. Using the DERS, they showed that emotion dysregulation and impulsivity contributed to excessive viewing. While emotion dysregulation involves trouble controlling internal states, impulsivity reflects poor behavioural inhibition—together, they point to executive dysfunction as a driver of compulsive media use (Kircaburun et al., 2024).

##### Study 5: TikTok Use, Self-Control, and Phubbing

Across two studies, researchers found that increased time spent on TikTok was significantly associated with lower self-control and greater phubbing behaviour—the tendency to ignore others in favour of one’s phone—while this pattern was not observed with Instagram Reels or YouTube Shorts. Self-control mediated the link between TikTok use and phubbing, and attention-shifting difficulties further explained users’ reduced ability to disengage from the app (David & Roberts, 2024).

##### Study 6: Executive Breakdown via Attention Control Network Analysis

Using network analysis, researchers examined the links between boredom proneness, attention control, and problematic short video use (PSVU). They found that difficulties with executive attention, particularly inattention and cognitive conflict (e.g., shifting, sustaining, resisting distraction), were central to the network, suggesting that poor attention regulation may predict and result from excessive SFV use. These findings support a feedback loop in which impaired executive control leads to boredom-driven media use, further depleting attention resources and reinforcing compulsive engagement (Zhou et al., 2024).

##### Synthesis and Relevance

These studies present a converging narrative: executive control difficulties are both measurable and subjectively experienced by SFV users daily. These findings deepen our understanding of the behavioural ecology of executive dysfunction, suggesting that SFV may reduce users’ ability to maintain control over their thoughts, emotions, and behaviours, especially in fast-paced, overstimulating digital environments.

| Study | Conclusion |
| --- | --- |
| Kircaburun et al., 2024 | Emotion dysregulation and impulsivity contribute to problematic mukbang viewing as a coping mechanism. |
| David & Roberts, 2024 | TikTok use, unlike other SFV platforms, is linked to increased phubbing through reduced self-control and attention-shifting difficulties |
| Zhou et al., 2024 | Inattention and conflict are key drivers in the feedback loop between poor executive control and problematic SFV use |

##### Overall Conclusion of Executive Functions-related Results

The reviewed studies show that SFV use disrupts executive functioning across neural, behavioural, and subjective domains. Experimental and neurophysiological research—using behavioural tasks, EEG, and fMRI—revealed subtle impairments in inhibitory control, conflict resolution, and emotional regulation, alongside reduced engagement of top-down executive systems in response to reward-driven cues. Complementing these results, self-report studies indicate that users experience diminished self-control, difficulty shifting attention, and heightened emotional impulsivity in everyday life, particularly when SFV is used to cope with boredom or distress. Together, these findings point to a reinforcing feedback loop: weakened executive control contributes to compulsive SFV use, which in turn further undermines cognitive regulation. This convergence of neural and subjective evidence underscores the potential for SFV platforms to erode sustained attention, behavioural inhibition, and emotional self-regulation over time.

| Theme | Number of Studies | Example Study |
| --- | --- | --- |
| Inhibitory control deficits | 3 | Jones et al., Su et al., Zhou |
| Emotion regulation issues | 2 | Su et al., Kircaburun et al. |
| Impulsivity | 3 | Nigbur et al., Su et al., Kircaburun |
| Neural reward dominance | 1 | Su et al. (VTA activation) |
| Executive attention shift/downregulation | 2 | Yan et al., David & Roberts |

#### 3.1.2. Critical Thinking

Two studies examined the impact of short-form media use on critical thinking and related metacognitive processes. While cross-sectional and survey-based, they offer unique insights into how digital content environments influence reflective thought, analytical reasoning, and cognitive self-awareness.

##### Study 2: Critical Thinking and Reasoning via CTAS and CRA

Fabio and Iaconis (2024) examined whether critical thinking protects against social media addiction among TikTok and Instagram users. Using the Critical Thinking Attitude Scale (CTAS) and the Critical Reasoning Assessment (CRA), they found that higher critical thinking scores were significantly associated with lower levels of addiction. Escapism partially mediated this relationship, indicating that individuals with lower essential abilities of thinking are more likely to engage in SFV social media as a coping mechanism, increasing their risk of addiction.

##### Study 8: Metacognitive Beliefs and Problematic Instagram Use

Albery et al. (2024) explored how Instagram-specific metacognitive beliefs and desire thinking predict problematic Instagram use. They found that negative metacognitions—such as beliefs about the uncontrollability and danger of one’s Instagram-related thoughts— alongside verbal perseveration (persistent self-talk about using the platform) significantly predicted compulsive use. In contrast, identity centrality (how important Instagram is to one’s self-concept) and imaginal prefiguration (mentally visualising Instagram use) were stronger predictors of withdrawal symptoms. While not a direct measure of critical thinking, the findings underscore how poor metacognitive regulation and desire-based thinking may contribute to addictive patterns of SM use.

##### Synthesis and Relevance

Together, these studies suggest that weaker critical thinking and maladaptive metacognitive patterns are associated with PMSU.

Both studies suggest a broader cognitive profile among users who may struggle to pause, question, or redirect their engagement with digital content. This suggests that strengthening critical and metacognitive thinking may be a valuable target for digital mental well-being.

| Study | Conclusion |
| --- | --- |
| Fabio and Iaconis, 2024 | Higher critical thinking scores were associated with lower SM addiction, with escapism partially mediating this relationship. |
| Albery et al., 2024 | Negative metacognitions and desire thinking predicted compulsive Instagram use, while identity centrality drove withdrawal symptoms. |

#### 2.1.3. Attention

##### Experimental Studies

Three studies in the current review employed objective, experimental tasks to assess different dimensions of attention among users of SFV platforms. Each of these studies utilised well-established cognitive paradigms—namely, the anti-saccade task, Attention Network Task (ANT), and Stroop task with eye tracking—providing insight into how SFV use might disrupt specific attentional processes.

##### Study (1): Inhibitory Control via Anti-Saccade Task

As reviewed in our examination of executive function, Jones et al. (2024) examined the relationship between SM use, inhibitory attentional control, and psychological distress using objective app usage data and an Anti-saccade task. Contrary to expectations, they found no negative association between SMU and attentional control; instead, a slight positive correlation emerged, particularly with the use of TikTok. Through the lenses of attention, this suggests that frequent engagement with dynamic visual content may modestly relate to enhanced inhibitory control, challenging the assumption that SMU inherently impairs attention regulation. However, these associations were minimal and should be interpreted cautiously.

##### Study (4): Executive and Alerting Attention via the Attention Network Task (ANT)

Similarly, echoing earlier results, Yan et al. (2024) investigated the impact of SFV use on attentional networks using the Attention Network Task (ANT) and EEG recordings.

Behavioural measures showed no deficits in alerting, orienting, or executive control; however, neurophysiological data revealed a significant negative correlation between SFV addiction tendency and theta power in the Frontal Cortex during executive control tasks, suggesting heavy SFV users may exhibit reduced neural engagement in cognitive control processes, even if behavioural performance appears intact.

##### Study (12): Attentional Disengagement and Focus via Stroop and Eye Tracking

Chen et al. (2023) investigated how addiction to SFV affects attention using both a video-watching task and a Stroop task with eye-tracking technology. Addicted users showed lower concentration, more distractions, and reduced interest when watching longer videos. Eye-tracking data revealed more fixations and shorter fixation durations, suggesting fragmented and unstable attention. During the Stroop task, participants exhibited slower reaction times, lower accuracy, and longer fixations, particularly under incongruent conditions, suggesting impaired attentional control and increased difficulty in filtering out distractions. These behavioural and gaze patterns reflect reduced sustained and executive attention among addicted users.

##### Synthesis and Relevance

Together, these three studies provide evidence that SFV use may impair distinct but interrelated domains of attention:

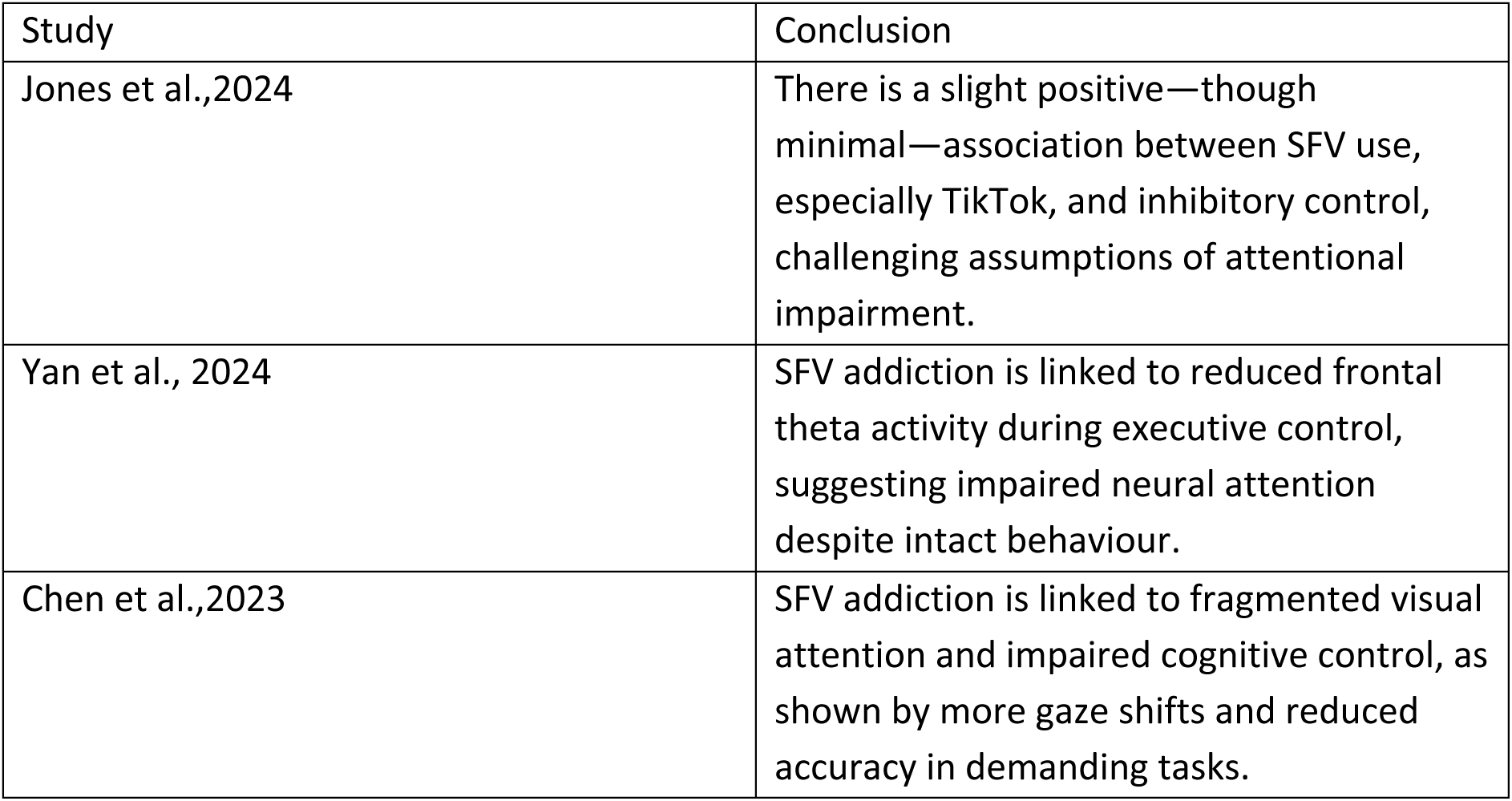

##### Studies Using Scales and Self-Reported Measures

While experimental tasks offer insight into discrete attention, two studies have adopted survey-based methods, and one employed a mixed-methods approach to assess subjective disruptions through psychometric and self-reporting tools.

##### Study 6: Network Analysis of Boredom, Attention, and Cognitive Overload

Previously reviewed, Zhou et al. (2024) used network analysis to explore the interaction between boredom, attention control, and problematic SVF use. Results revealed that boredom proneness was the most central psychological trait driving both attentional dysfunction and problematic SVF use. Consistent with what is starting to appear as a recurrent theme, the results suggest that individuals with poor attention control may become chronically bored, leading to compulsive SFV use as a coping strategy, reinforcing the loop of digital distraction and cognitive overload.

##### Study 7: Real-World Attention Deficits in Mixed-Methods Design

Lin et al. (2024) conducted a mixed-method study combining an online survey with an 18-day field experiment to assess how SFV consumption impacts sustained attention. In the survey phase, higher daily SFV use was significantly associated with more significant commission errors and variability in response times on the Sustained Attention to Response Task (SART), indicating behavioural deficits in vigilance and attentional stability. The field experiment manipulated daily video viewing (increasing or decreasing short-vs. long-form content) and revealed that increased SFV consumption led to higher self-reported attentional lapses (ARCES). However, task-based improvements suggested potential learning effects. No significant changes were found in perceived attention via the Mindful Attention Awareness Scale – Lapse Only (MAAS-LO), highlighting a disconnect between subjective and objective attention measures. Overall, the findings suggest that frequent SFV consumption may undermine both perceived and actual sustained attention in everyday contexts.

##### Study 14: Self-Reported Attention Using a Custom Checker

Srivastava (2023) explored how SFV exposure affects sustained attention and information comprehension through a self-report custom-designed survey and interview that revealed a negative correlation between time spent on SFV platforms and perceived ability to sustain attention, with heavy users reporting a 35% reduction in attention span and more difficulty processing complex digital texts and videos. Participants described how habitual SFV consumption conditioned them to expect rapid content delivery, making them restless and distracted when faced with longer, cognitively demanding material. These self-perceived difficulties included maintaining focus, following detailed arguments, and retaining information.

##### Synthesis and Relevance

Together, these three studies demonstrate that SFV addiction impairs both sustained and executive attention, not only in controlled tasks but also in everyday cognitive functioning.

| Study | Conclusion |
| --- | --- |
| Zhou et al., 2024 | Boredom proneness and low attention control jointly drive problematic SFV, reinforcing a cycle of distraction and cognitive overload. |
| Lin et al., 2024 | High SFV consumption is linked to behavioural and perceived deficits in sustained attention. |
| Srivastava, 2023 | High SFV exposure is linked to reduced self-perceived sustained attention and comprehension, with users reporting restlessness and cognitive fatigue when engaging with longer content. |

##### Overall Conclusion of Attention-related Results and Findings

Evidence suggests that SFV use is consistently associated with impairments in sustained and executive attention across six studies. Experimental results indicated disruptions in domains requiring sustained mental focus, interference suppression, and visual stability. Self-reporting revealed that habitual SFV consumption undermines users’ perceived attentional capacity through mind wandering, difficulty processing complex content, and a growing dependence on rapid content shifts. These results suggest that SFVs may condition users toward shallow cognitive engagement and attentional volatility.

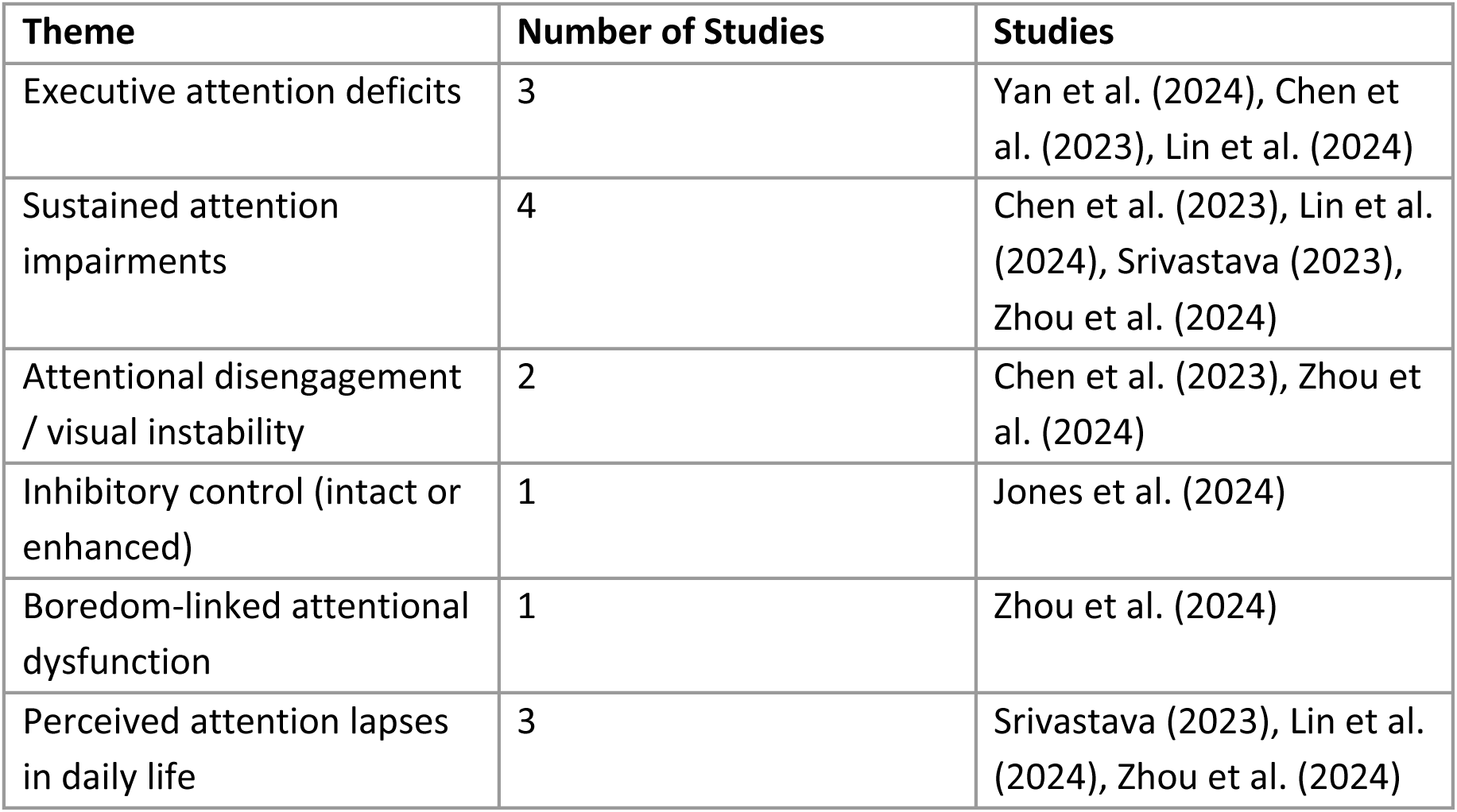

| Theme | Number of Studies | Studies |
| --- | --- | --- |
| Executive attention deficits | 3 | Yan et al. (2024), Chen et al. (2023), Lin et al. (2024) |
| Sustained attention impairments | 4 | Chen et al. (2023), Lin et al. (2024), Srivastava (2023), Zhou et al. (2024) |
| Attentional disengagement / visual instability | 2 | Chen et al. (2023), Zhou et al. (2024) |
| Inhibitory control (intact or enhanced) | 1 | Jones et al. (2024) |
| Boredom-linked attentional dysfunction | 1 | Zhou et al. (2024) |
| Perceived attention lapses in daily life | 3 | Srivastava (2023), Lin et al. (2024), Zhou et al. (2024) |

#### 3.1.4. Memory

Two studies investigated the impact of engagement with SFV on memory, specifically examining both long-term (prospective and retrospective) and short-term memory functions. Although differing in design, both studies indicate cognitive disruptions.

##### Study 12: Memory and Academic Impairment via Self-Report

Xia et al. (2023) conducted a large-scale survey to investigate how SFV affects academic functioning through the mediating role of memory and mood. Findings showed that students who spent more time on SFVs had significantly lower scores on memory measures, particularly in prospective memory—the ability to remember to carry out intended actions in the future. This memory decline was accompanied by increased anxiety and depression. It served as a significant mediator in the relationship between SFV use and academic delay of gratification, suggesting that SFV-induced memory impairments may indirectly weaken goal-oriented behaviour. This study positions prospective memory as a vital cognitive mechanism linking SFV overuse to poor academic self-regulation.

##### Study 17: Short-Term Memory Recall in an Experimental Setting

Spence et al. (2020) examined the effects of Instagram use during versus after the presentation of auditory information on short-term memory in college students. Using the Logical Memory Immediate Recall (LM I) subtest from the Wechsler Memory Scale IV, participants were assigned to one of three groups: no Instagram use, Instagram during story presentation, or Instagram after. Results showed that using Instagram during the material presentation significantly reduced memory performance compared to the control group while using Instagram after the presentation did not produce a significant difference.

Neither the number nor the type of Instagram feed topics moderated memory performance. These findings underscore the cognitive cost of media multitasking, particularly when SM competes with real-time learning, and suggest that timing, not just the presence of distraction, plays a critical role in short-term memory encoding.

##### Synthesis and Relevance

Despite differing in design, both Xia et al. (2023) and Spence et al. (2020) provide converging evidence that SFV harms memory.

| Study | Reference |
| --- | --- |
| Xia et al., 2023 | Excessive SFV use is linked to impaired prospective memory, which mediates reduced academic self-control through elevated anxiety and depression. |
| Spence et al., 2020 | Scrolling Instagram while receiving new information significantly impairs short-term memory, highlighting the attentional cost of media multitasking during learning. |

#### 3.1.5. Emotional Regulation

Two studies investigated the impact of engagement with SFV platforms on emotional regulation. Both studies employed survey-based methods but focused on distinct emotional and behavioural outcomes. Together, they provide insight into how emotionally immersive and impulsivity-driven interactions with SFV content may serve as maladaptive coping strategies, potentially undermining users’ broader emotional regulation capacities.

##### Study 3: Emotion Regulation Difficulties and Psychological Distress

Kircaburun et al. (2024), as previously discussed, examined how emotion regulation difficulties (ERDs) and psychological distress contribute to problematic mukbang watching (PMW). Findings showed that higher ERDs were both directly and indirectly linked to PMW through increased anxiety and positive urgency—a facet of impulsivity. Although all impulsivity traits correlated with PMW, only positive urgency remained significant in the complete model, suggesting that some individuals may use emotionally charged online video content as a maladaptive coping strategy in response to overwhelming emotions.

Notably, anxiety was a significant mediator, pointing to the specific emotional pathways by which SFV-type content consumption may become compulsive. The study provides evidence that emotion dysregulation, particularly under emotional reactivity and poor impulse control conditions, plays a key role in problematic engagement with SFV.

##### Study 5: Self-Control, Emotional Coping, and Phubbing

David and Roberts (2024) investigated how different flow states experienced while using TikTok and Instagram relate to psychological well-being, including mind-wandering, addiction, and emotional distress. Their findings revealed that users experiencing high levels of telepresence (deep immersion in the platform) reported significantly greater anxiety, depression, and compulsive usage, especially among those with high overall flow. These effects were more substantial for TikTok users, who reported higher levels of time distortion and enjoyment, reinforcing that SFV platforms may encourage users to disengage from reality and emotionally overindulge in escapist content. The study supports the notion that telepresence is the key emotional mechanism linking flow states with adverse psychological outcomes, acting as a maladaptive emotional regulation strategy by displacing real-world coping resources and social interactions.

##### Synthesis and Relevance

Together, these studies indicate that high engagement with SFV content may impair emotional regulation by reducing users’ ability to manage impulses and stress:

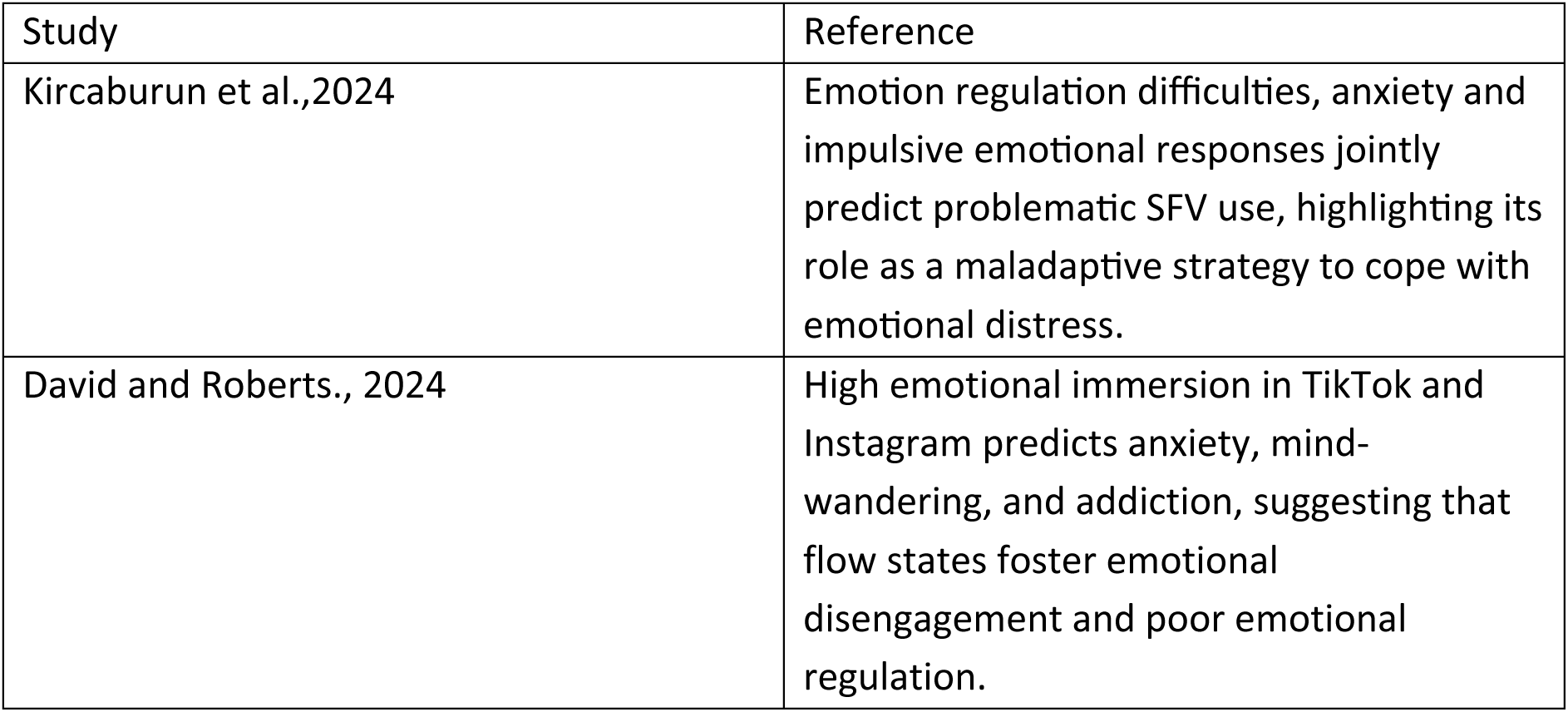

| Study | Reference |
| --- | --- |
| Kircaburun et al.,2024 | Emotion regulation difficulties, anxiety and impulsive emotional responses jointly predict problematic SFV use, highlighting its role as a maladaptive strategy to cope with emotional distress. |
| David and Roberts., 2024 | High emotional immersion in TikTok and Instagram predicts anxiety, mind-wandering, and addiction, suggesting that flow states foster emotional disengagement and poor emotional regulation. |

##### Integrative Summary of Cognitive Conclusions

The reviewed studies present a complex picture of how SFV consumption influences cognitive mechanisms among Gen Z. Overall, frequent SFV use is consistently linked to attentional fragmentation, diminished executive control, and increased impulsivity.

Neuroimaging data support this, revealing reduced activation in frontal regions responsible for cognitive regulation in individuals who use high amounts. Behavioural studies employing tasks such as SART and ANT highlight reduced sustained attention and variable performance consistency among heavy SFV users. Self-report measures corroborate these findings, with users reporting reduced attention span, impaired information retention, and a heightened reliance on rapid content delivery. Importantly, a recurrent theme across studies is the mismatch between preserved behavioural task performance and underlying neurocognitive strain, suggesting that SFV use may mask deficits until cognitive load intensifies.

Overall, SFV consumption appears to condition attentional habits toward novelty and speed, challenging the capacity for deep, sustained focus. These patterns raise concerns about long-term effects on cognitive flexibility, learning, and emotional self-regulation, especially in developmental contexts. While more longitudinal research is needed, current findings indicate that the cognitive impacts of SFV are not merely transient or superficial—they may represent enduring shifts in how attention and executive functions operate in digital natives.

### 3.2. Mental Health

Building on the cognitive dimensions explored, this section shifts focus to the psychological outcomes associated with SFV consumption among Gen Z. Drawing on some of the previously mentioned studies and more, cross-sectional, experimental, and scale-based studies examined the relationship between SFV use and mental health indicators such as depression, anxiety, well-being, and patterns of problematic engagement.

#### 3.2.1. Depression

Three studies explored the relationship between SM and depressive symptoms, focusing on the spectrum of SFV, mukbang watching, and online gaming. Using cross-sectional surveys, these studies identified distinct emotional and behavioural mechanisms—such as anxiety, impulsivity, and academic disengagement—that may indirectly contribute to depression.

They highlight that not all screen time is equal, and the type, purpose, and emotional context play a role in shaping mental health outcomes.

##### Study 1: Psychological Distress in General Social Media Use

Jones et al. (2024) examined attentional control and depression using the Depression Anxiety Stress Scales DASS-21. The study found no significant association between overall time spent on SM and depressive symptoms and only a minimal positive association between total SMU and anxiety. Interestingly, platform-specific analysis revealed that Facebook use—but not TikTok or Instagram—was weakly associated with increased psychological distress, suggesting that the nature of content or interaction style on specific platforms may matter more than total usage time. These findings challenge common assumptions based on self-reported data and highlight that actual time spent on SFV platforms may not directly contribute to depressive symptoms. Instead, how individuals interact with the content and the platform-specific dynamics may better explain mental health risks.

##### Study 3: Emotional Dysregulation and Depression in Mukbang Viewing

Kircaburun et al. (2024) investigated whether emotion regulation difficulties (ERDs), impulsivity, and psychological distress—including depression—predict problematic mukbang watching (PMW). While depression showed a weak positive correlation with PMW, it was not a significant predictor in the final path model. Instead, ERDs were directly and indirectly associated with PMW, primarily through anxiety and the impulsivity trait of positive urgency. These findings suggest that PMW may function as a maladaptive coping strategy, particularly for individuals who struggle with anxiety and emotional reactivity, rather than depressive symptoms specifically.

##### Study 11: Depression and Procrastination in SFV and Gaming Users

Xia et al. (2023) investigated the impact of excessive SFV and online gaming use on college students’ mental health and academic performance. They found that higher daily usage was significantly associated with increased depression scores, as measured by the PHQ-9, along with declines in prospective memory and academic delay of gratification (ADOG). Path analysis showed that depression and impaired prospective memory served as key mediators, suggesting that excessive SFV use may indirectly reduce students’ academic motivation and self-regulation by first undermining emotional and cognitive stability.

##### Synthesis and Relevance

Together, these studies suggest that depression is not directly tied to SFV use but may emerge indirectly through emotional or cognitive pathways such as anxiety or academic disengagement. The context and purpose of use appear more influential than screen time alone.

| Study | Conclusion |
| --- | --- |
| Jones et al., 2024 | There is no link between SFV and depression, but weak platform-specific associations suggest that how and where users engage may influence psychological distress. |
| Kircaburun et al., 2024 | Anxiety—but not depression—mediated the link between emotion dysregulation and PMW, highlighting anxiety-driven emotional coping. |
| Xia et al., 2023 | Depression mediates the relationship between excessive SFV and reduced academic motivation. |

#### 3.2.2. Anxiety

Two studies examined the relationship between anxiety symptoms and patterns of SFV, highlighting distinct mechanisms through which digital engagement may contribute to heightened anxiety.

##### Study 1: General Psychological Distress Including Anxiety

Jones et al. (2024) assessed anxiety using the DASS-21 and found no significant association between total SM and anxiety symptoms. However, subtle elevations in distress were observed among users who engaged passively and frequently. This pattern suggests that unstructured and repetitive exposure to emotionally stimulating content may contribute to low-level anxiety, possibly through cognitive overstimulation and impaired emotional recovery mechanisms.

##### Study 3: Anxiety/Stress and Emotional Dysregulation

In contrast, Kircaburun et al. (2024) identified anxiety—not depression or stress—as the primary emotional mediator linking emotion regulation difficulties to PMW, a niche form of SFV use. Individuals with high emotional reactivity and poor impulse control were more likely to use mukbang content as a maladaptive coping strategy, relying on its sensory stimulation to escape anxious thoughts. The study highlights how emotionally immersive digital media may reinforce anxiety through avoidant coping loops.

##### Synthesis and Relevance

Together, these studies suggest that anxiety may be subtly intensified by habitual SFV use, particularly when content is consumed passively or as a form of emotional escape. While general use shows only weak associations, anxiety appears more pronounced when emotional dysregulation and avoidant coping are involved.

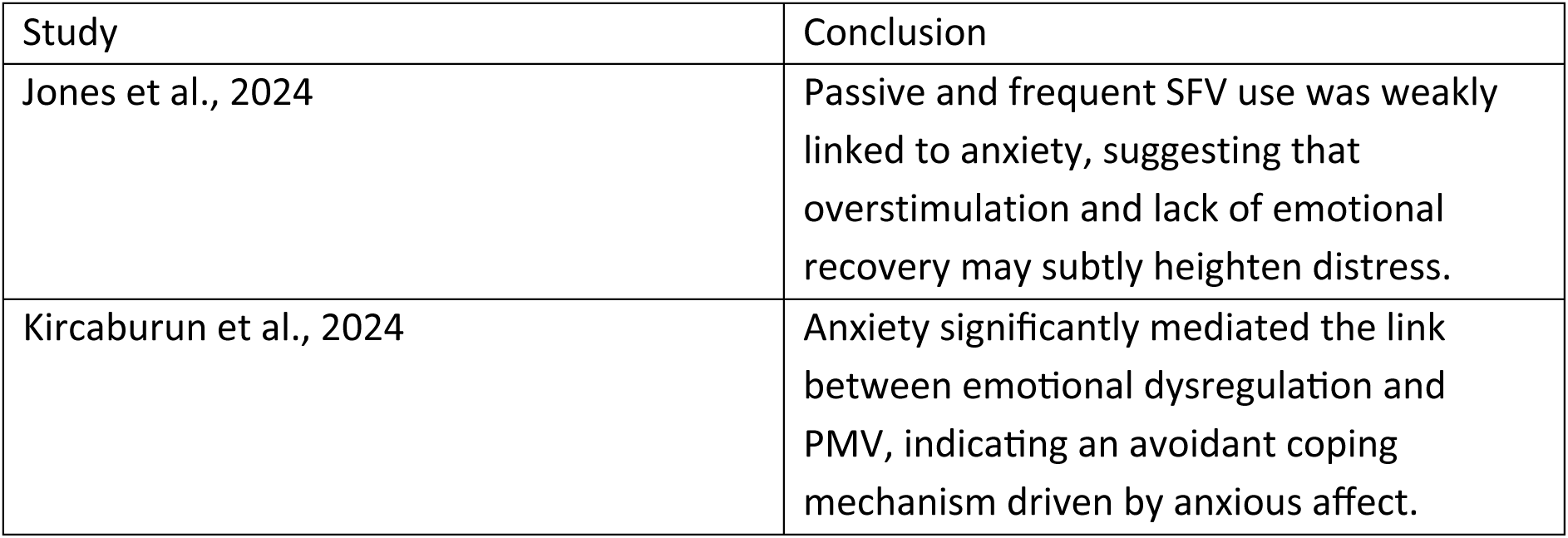

| Study | Conclusion |
| --- | --- |
| Jones et al., 2024 | Passive and frequent SFV use was weakly linked to anxiety, suggesting that overstimulation and lack of emotional recovery may subtly heighten distress. |
| Kircaburun et al., 2024 | Anxiety significantly mediated the link between emotional dysregulation and PMV, indicating an avoidant coping mechanism driven by anxious affect. |

*3.2.3 General well-being, Life Satisfaction and Mood.*

#### 3.2.3 General well-being, Life Satisfaction and Mood

The reviewed studies on how SFV affects mood, life satisfaction, and overall psychological well-being revealed both adverse outcomes, such as emotional fatigue and digital dependency, and potential benefits, such as enhanced mood and emotional receptivity, further underscoring the nuanced effects of SFV engagement on emotional health.

##### Study 9: Information Overload and Life Satisfaction

Chung et al. (2023) investigated how perceived information overload from social media video platforms affects well-being. Their findings showed that information overload was significantly linked to life dissatisfaction. This effect was fully mediated by SFV fatigue and maladaptive coping, with a partial mediation by life dissatisfaction itself. The results highlight that digital overwhelm—not necessarily the content itself—can erode psychological well-being by draining cognitive and emotional resources, leading to disengagement and a reduction in life satisfaction.

##### Study 10: Flow States and Affective Engagement

Roberts and David (2023) explored how different flow states—such as enjoyment, time distortion, and especially telepresence—experienced on TikTok and Instagram relate to psychological well-being. They found that higher levels of telepresence were strongly associated with increased depression and anxiety, as well as greater addiction, mind-wandering, and FOMO. TikTok users reported more intense flow experiences, suggesting that immersive, emotionally engaging SFV content can displace real-world coping and connection, undermining emotional balance and long-term well-being.

##### Study 15: Mood Modulation

Nigbur and Ullsperger (2020) used EEG to investigate the effects of humorous SFV on mood and cognitive processing. Their results showed that positive mood induction via funny SFV enhanced neural markers of emotional receptivity and performance monitoring. These findings suggest that brief exposure to uplifting digital content can improve mood and promote adaptive emotional engagement, challenging the view that all SFV consumption is detrimental.

##### Synthesis and Relevance

While information overload and immersive flow states can undermine life satisfaction and emotional balance, deliberate exposure to positive, mood-enhancing content may offer short-term emotional benefits. This contrast suggests that the psychological effects of SFV use are not universally harmful but depend heavily on intent, content type, and user regulation.

| Study | Conclusion |
| --- | --- |
| Chung et al., 2023 | Information overload from SFV platforms reduces life satisfaction by fostering digital fatigue and maladaptive coping. |
| Roberts and David, 2023 | Immersive flow states predict higher anxiety, depression, and digital dependency, pointing to emotional over-absorption and poor coping. |
| Nigbur and Ullsperger, 2020 | Humorous SFV clips boosted positive mood and emotional receptivity. |

#### 3.2.4. Addiction and Problematic Use

Drawing on evidence from self-report, behavioural tasks, eye-tracking, EEG, and fMRI, the reviewed studies reveal how SFV addiction is associated with reduced executive functioning, impaired attentional control, maladaptive metacognitive beliefs, and altered brain activity in reward and self-regulation circuits. Together, these findings suggest that compulsive SFV engagement emerges not solely from user habits but from an interaction between internal vulnerabilities and platform-driven reinforcement, raising important questions about digital media design and mental autonomy.

##### Study 4: Neural Correlates of SFV Addiction

Yan et al. (2024) investigated how tendencies toward SFV addiction impact cognitive control, finding that higher addiction scores were significantly linked to reduced executive functioning. These effects were independent of anxiety and depression and strongly correlated with lower self-control.

##### Study 8: Metacognitive Roots of Problematic Use

Albery et al. (2024) examined how metacognitive beliefs and desire thinking contribute to problematic Instagram use. They found that negative metacognitive beliefs, verbal perseveration, imaginal prefiguration, and identity centrality predicted compulsive use and withdrawal, suggesting that problematic SFV engagement is shaped more by internal cognitive and identity-based processes than by external platform design, particularly when users rely on the app to manage distressing thoughts or reinforce their self-concept.

##### Study 12: Eye-Tracking Indicators of SFV Addiction

Chen et al. (2023) examined attentional control in users with SFV addiction. Addicted users showed more gaze shifts, shorter fixation durations, and more excellent distraction while watching videos, along with poorer accuracy and longer response times during the Stroop task. These results indicate that SFV addiction is associated with reduced sustained attention and impaired interference control, both during and after video exposure. The study provides objective, behavioural evidence that addictive SFV use fragments attentional stability and may undermine cognitive regulation.

##### Study 17: fMRI and Algorithm-Driven Compulsivity

Su et al. (2021) examined how TikTok’s personalised recommendation algorithm contributes to problematic SFV use, combining survey data with fMRI analysis. Participants who viewed algorithm-curated videos exhibited heightened activation in reward-related brain regions and reduced connectivity in self-control-related areas. Problematic use was also negatively correlated with self-control scores, suggesting that TikTok’s algorithm may capture attention and actively reinforce compulsive engagement by modulating the brain’s reward and regulatory systems.

##### Synthesis and Relevance

Together, these studies highlight the cognitive and neurobiological underpinnings of SFV addiction and compulsive use. Findings indicate that problematic engagement is not only associated with reduced executive function and attentional control but is also influenced by maladaptive metacognition and desire-based rumination. At the neural level, algorithm-driven reward stimulation and weakened self-regulation circuits appear to reinforce addictive patterns, suggesting that both internal vulnerabilities and external design features contribute to sustained, dysregulated SFV use.

| Study | Conclusion |
| --- | --- |
| Yan et al., 2024 | SFV addiction is linked to reduced executive functioning. |
| Albery et al., 2024 | Negative metacognitions and desire thinking predict compulsive SFV use. |
| Chen et al., 2023 | SFV addiction impairs attentional control and increases distractibility. |
| Su et al., 2021 | TikTok’s algorithm drives compulsive use by activating reward circuits and dampening self-control networks |

#### 3.2.5. Escapism, Boredom, Flow, Phubbing, Maladaptive Coping

The following studies explore the psychological drivers behind SFV use, focusing on how escapism, boredom, and digital immersion contribute to compulsive engagement. Patterns of emotional avoidance, impaired self-regulation, and maladaptive coping emerge as key risk factors for declining well-being.

##### Study 2: Escapism, Flow, and Low Critical Thinking

Fabio and Iaconis’s (2024) findings revealed that escapism was a strong predictor of addictive engagement on TikTok and Instagram. This effect was amplified by immersive flow states and a sense of community belonging, which increased time spent on the platforms.

Individuals with lower critical thinking disposition and reasoning skills were more prone to such maladaptive coping, using SFVs to bypass emotional discomfort rather than reflectively process it. Mediation analyses confirmed that escapism partially explained the link between low critical thinking and higher addiction risk, suggesting that insufficient reflective capacity may predispose users to emotionally driven, compulsive digital habits.

##### Study 5: Phubbing and Emotional Avoidance

David and Roberts (2024) highlighted that users spending more time on platforms like TikTok and Instagram were significantly more likely to phub others, displaying lower self-control and a reduced capacity for face-to-face emotional engagement. They suggested that the immersive flow states (telepresence and time distortion) triggered by SFVs foster compulsive scrolling behaviours that can displace meaningful offline connections. These findings support the notion that SFVs can become tools for emotional avoidance and social disconnection, reinforcing maladaptive digital coping mechanisms rather than promoting interpersonal or psychological resilience.

##### Study 6: Boredom and Compulsive Engagement

Zhou et al. (2024) found that boredom proneness is the central trait, linking low arousal states with attention disengagement and compulsive video consumption. The inattention facet had the highest centrality across the network, suggesting that difficulty sustaining focus may drive individuals to seek stimulation through SFVs. The study supported the boredom feedback model, proposing that low-attentional control users are more susceptible to a regulatory loop in which SFV provide short-term relief from boredom but worsens attentional fatigue over time. This dynamic reinforces compulsive digital engagement as a maladaptive strategy for coping with both under- and over-stimulation.

##### Study 10: Maladaptive Coping and Information Overload

Chung et al. (2023) focused on maladaptive coping in response to perceived information overload from SFV platforms. Users who felt overwhelmed by the volume and pace of content were more likely to use SFV to escape stress but reported lower life satisfaction and higher emotional fatigue. This underscores how cognitive overload may foster dysfunctional coping through digital media, further reducing psychological well-being.

Chung et al. (2023) found that Chinese young adults who felt overwhelmed by the constant flow of notifications and content on SFV apps were likelier to adopt maladaptive coping strategies, such as emotional avoidance or disengagement. These coping styles were associated with increased emotional fatigue and life dissatisfaction. Mediation analyses confirmed that maladaptive coping and fatigue fully mediated the relationship between perceived overload and the intention to discontinue SFV use. This suggests that cognitive overload does not simply reduce user engagement through fatigue—it also compromises emotional regulation and overall psychological resilience.

##### Synthesis and Relevance

These studies highlight that problematic SFV use is less about screen time and more about the underlying psychological motives driving engagement. Emotional avoidance, boredom regulation, and cognitive overwhelm emerge as central triggers for compulsive SFV consumption. This body of research underscores that the motivational architecture of SFV use, shaped by unmet emotional and mental needs, is a more potent predictor of maladaptive outcomes than usage duration alone.

|  |  |
| --- | --- |
| Study | Conclusion |
| Fabio and Iaconis, 2024 | Lower critical thinking ability increases susceptibility to escapist and emotionally immersive SFV use, |
| David and Roberts 2024 | High SFV use is linked to phubbing and emotional withdrawal. |
| Zhou et al., 2024 | Boredom proneness and low attention control significantly predict compulsive SFV use. |
| Chung et al., 2023 | Perceived information overload on SFV platforms fosters maladaptive coping, emotional fatigue, and life dissatisfaction. |

### 3.3. Overall Conclusion

This synthesis reveals that SFV use exerts a multifaceted influence on both cognitive and mental health functioning among Gen Z. Rather than viewing these domains independently, the evidence highlights their reciprocal relationship: diminished executive control, attentional instability, and memory impairments appear to heighten emotional reactivity, impulsivity, and anxiety. In turn, these psychological vulnerabilities promote patterns of digital coping and compulsive use that further strain cognitive systems, particularly those involved in regulation and sustained engagement.

**Figure 4.**
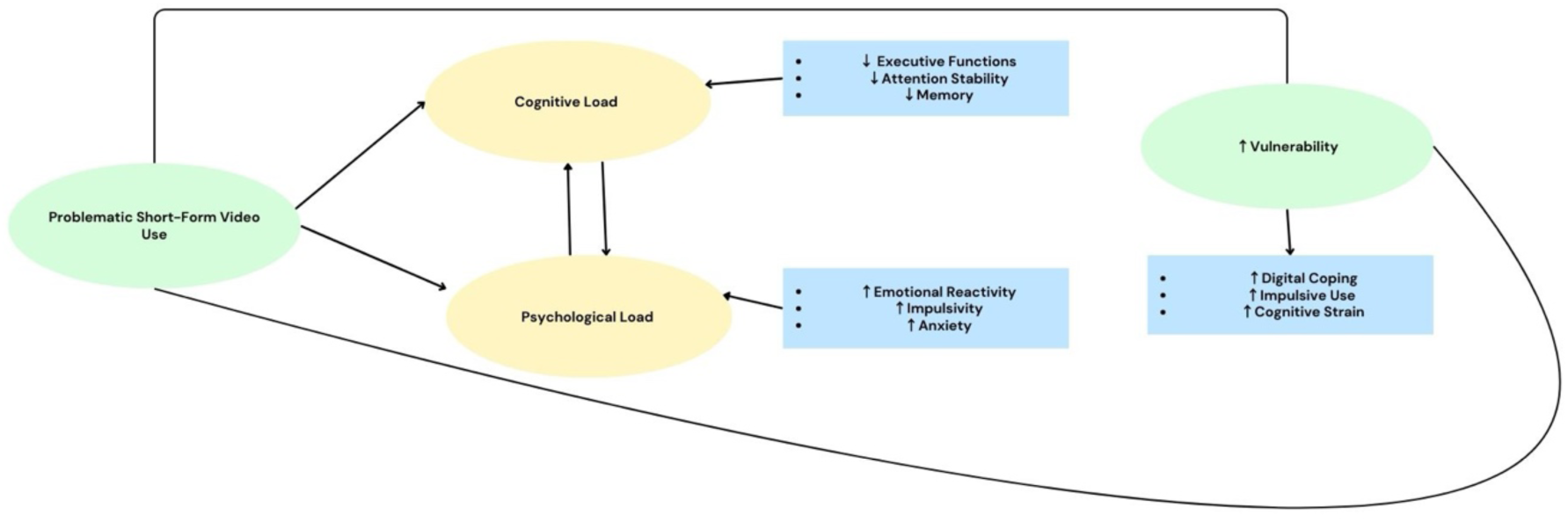
Interactive Effects of Cognitive and Psychological Load in Problematic SFV Consumption

Cognitively, neurophysiological studies indicate a shift from goal-directed to stimulus-driven processing, as evidenced by altered activation of control and reward circuits. Behavioural and self-report studies reinforce this trend, revealing difficulties with impulse regulation, attentional disengagement, and emotion-driven decision-making. Psychologically, SFV engagement is linked to a spectrum of mental health outcomes, including increased anxiety, emotional dysregulation, and digital dependency. However, these effects are neither uniform nor universally negative. In some contexts, brief exposure to humorous or emotionally uplifting SFV content can transiently improve mood and emotional receptivity. Yet, the convergence of flow states, escapism, boredom regulation, and maladaptive coping suggests that SFV platforms serve as a mechanism for emotional avoidance rather than resilience-building for many users, especially those with poor self-regulation.

The synthesis suggests that problematic SFV use is not merely a matter of screen time but is shaped by internal cognitive mechanisms, emotional dispositions, and platform design features. These factors form a reinforcing cycle in which users are drawn to SFVs to manage distress but may ultimately experience reduced self-control, attentional fatigue, and impaired psychological well-being.

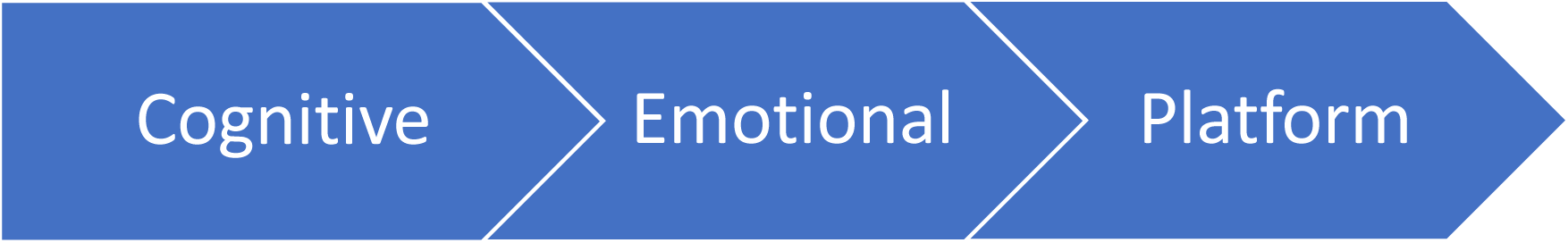

## 4. Discussion

### 4.1. Overview & Aims Recap

The review aimed to assess how SFV influences specific cognitive mechanisms and mental health outcome variables among Gen Z. Through a comprehensive search and critical synthesis of peer-reviewed studies published between 2014 and 2024, this work provides a consolidated understanding of the empirical landscape and highlights vital gaps in the literature.

### 4.2. Summary of Key Findings

Findings from the included studies indicate a mixed and complex picture. Overall, frequent SFV use was associated with attentional fragmentation, reduced sustained focus, and impaired working memory. The mental health impact of SFV use was inconsistent, with some studies reporting increased stress, anxiety, and compulsive behaviours, while others suggested mood enhancement or short-term emotional relief.

### 4.3. Integration of Results with Broader Theories

The results align with and extend broader research in digital media, cognitive science, and neuropsychology, offering a systems-level understanding of SFV effects on Gen Z.

#### Executive Control and Neurofunctional Shifts

SFV use appears to impair executive control, particularly attention regulation and inhibitory processes. These findings align with research which provides neuroimaging evidence of disrupted connectivity in networks governing executive function, salience, and reward processing—namely, the prefrontal cortex and striatum (Zhou & Wang, 2025). These changes suggest heightened reward sensitivity and compromise self-regulation, reinforcing behavioural observations of reduced attention stability. From a neurobiological lens, the DLPFC and ACC show decreased activation during SFV engagement (Su et al., 2021; Firth et al., 2019), indicating a shift toward reactive, stimulus-driven behaviour. These dynamics align with findings in digital addiction literature (Liu et al., 2025; Caponnetto et al., 2025), suggesting that persistent SFV use may cause lasting neural adaptations, especially in regions tied to top-down control and reward anticipation. Despite the prevalence of this comparison to substance-related and behavioural addictions, it is not officially classified as a mental disorder in current psychiatric manuals such as the DSM-5 or ICD-11 (Li, Liu, & Liu, 2024).

#### Working Memory Overload and Cognitive Load Theory

According to cognitive load theory (Sweller, 1988), the speed and density of SFV content can overwhelm the brain’s capacity to chunk and integrate information. This results in fragmented encoding, shallow processing, and reduced transfer to long-term memory.

Recent findings from Jiang and Ma (2024) support this by showing that even brief exposure to TikTok content reduces analytic thinking, promoting intuitive and low-effort processing.

#### Emotional Reactivity and Reward Loops

The pattern cycles of high arousal and gratification are consistent with incentive salience theory (Berridge & Robinson, 2003), which describes how repeated exposure to rewarding content increases the brain’s drive to “want” without a corresponding increase in genuine “liking.” This may explain the compulsive engagement and emotional dysregulation observed in users. Over time, this leads to hedonic adaptation—users require increasingly novel or extreme content to achieve the same emotional impact, mirroring behavioural addiction processes described by Caponnetto et al. (2025).

#### Boredom, Dopamine Loops, and Motivational Flattening

Recent studies suggest that digital media—especially SFV—can make people more bored, not less. Tam and Inzlicht (2024) found that overusing SM raises our need for constant stimulation and makes it harder to enjoy slower, more meaningful activities. This concept is linked to the idea of affective habituation and opportunity cost: when accustomed to fast-paced, exciting content, everyday life feels less engaging. Over time, this reduces motivation and narrows the ability to focus. SFVs, being quick and unpredictable, exacerbate this issue by constantly distracting the brain and contributing to inattention.

Together, these insights highlight the complex, multi-level mechanisms underlying SFV’s psycho-cognitive effects. The following section discusses the limitations of this review, including the need for caution when interpreting these findings.

### 4.4. Limitations of the Existing Literature and Review

The existing literature on SFV and its cognitive and psychological effects presents several key limitations. Most studies rely on cross-sectional designs, making it difficult to establish causality or determine the temporal order of observed associations, such as executive dysfunction or emotional dysregulation. Additionally, inconsistent operational definitions— particularly for constructs like attention, emotional regulation, and problematic use—limit comparability across studies and hinder synthesis efforts (Caponnetto et al., 2025).

Another persistent issue is the heavy reliance on self-report measures, which are susceptible to recall bias, social desirability bias, and subjective interpretation. These tools compromise ecological validity and reduce the accuracy of reported associations (McCarthy & Morina, 2020). Longitudinal and experimental designs remain scarce yet are essential for clarifying the cumulative and directional effects of SFV use over time.

A further limitation lies in the predominance of quantitative methods. While valuable, they often fail to capture the nuanced, lived experiences behind SFV engagement. First-person perspectives are largely absent, which limits our understanding of how digital habits relate to identity, coping and context. Scholars such as Livingstone and Sefton-Green (2016) and Pink et al. (2016) argue that digital media research should incorporate interviews, digital ethnography, or diary methods to reflect the complexity of everyday technology use more accurately. Sample diversity also poses a concern, as many studies draw from convenience samples of university students, which reduces generalisability. Moreover, even when neurobiological or behavioural models are used, they are rarely integrated with ecological or engagement data (e.g., algorithmic exposure or usage patterns), limiting a systems-level understanding of SFV effects. Finally, although SFV use is often framed in terms of behavioural addiction, it is not formally recognised as a clinical disorder in classifications such as the DSM-5 or ICD-11. This creates conceptual ambiguity, as addiction frameworks are applied without clear diagnostic benchmarks (Li, Liu, & Liu, 2024). As McCarthy and Morina (2020) note, this may contribute to overpathologising behaviours that are situational or developmentally normative.

### 4.5. Implications

The findings from this review offer several implications for research, clinical practice, public health, and digital policy.

For research, this synthesis highlights the need for more longitudinal and experimental designs to understand better the causal relationships between SFV use and cognitive-emotional outcomes. Establishing standardised definitions and validated cognitive metrics will also enhance comparability across studies. Moreover, integrating behavioural, neurobiological, and qualitative approaches may provide a more nuanced, mechanistic understanding of SFV’s effects. For clinical and educational practice, professionals working with Gen Z should be aware of SFV’s potential to aggravate attention difficulties, emotional dysregulation, and compulsive behaviours. Media use assessments may be especially relevant in cases presenting with anxiety or executive dysfunction. At the policy level, critical media literacy programmes and tailored screen-time guidance could help mitigate risks related to algorithm-driven overstimulation. These findings collectively point to the need for a deeper understanding of user perspectives—an aim best addressed through qualitative inquiry, which is introduced in the following section.

### 4.6. Linking to the Qualitative Study

This review serves as the foundation for a broader mixed-methods study. While it identified consistent associations between SFV use and cognitive-emotional effects, it also exposed a critical gap: how Gen Z users themselves perceive and make sense of these experiences.

Most existing studies overlook lived meaning-making. To address this, the next phase involves a qualitative study using semi-structured interviews with university students. It will explore perceived agency, attentional self-awareness, and emotional coping in relation to SFV use. Framed within a constructivist and abductive approach, this phase seeks to develop theory-informed constructs grounded in user experience. These insights will inform the design of a follow-up quantitative tool, enhancing its conceptual validity.

In this way, the qualitative phase bridges the empirical findings of the review with users’ subjective perspectives, advancing a more contextually grounded understanding of SFV’s cognitive and emotional impacts.

### 4.7. Conclusion

This review highlights the multifaceted cognitive and emotional implications of SFV use among Gen Z, with recurrent patterns of attentional disruption, executive dysfunction, and mental health vulnerability. Yet, the existing literature remains limited by cross-sectional designs, inconsistent measures, and an over-reliance on self-reported data. These methodological constraints underscore the need for interdisciplinary approaches that integrate both objective indicators and subjective accounts.

In response, the next study presents the methodological framework which seeks to examine how young users perceive and enact agency and control in relation to SFV use. By combining subjective experiences with objective cognitive and emotional metrics, the aim is to advance a more nuanced understanding of SFV’s impact on mental health and cognition.

## Supporting information

authors and affiliations updated.

## Data Availability

All data is included in supplementary files.

## Data Availability

All data is included in supplementary files.

## Appendix

### 7.1 Data Extraction – Full Table

See Supplementary File No. 1 – attached.

### 7.2. Author’s Details

See Supplementary File No. 2 – attached.

### 7.3. Ethical Approval

See Supplementary File No.3 - attached

