## Supplementary material for "The Impact of Short-Form Video Use on Cognitive and Mental Health Outcomes: A Systematic Review": authors and affiliations updated.

**Author Details and Contributions**

**Sara Arouch**

Affiliation: Centre of Intelligent Healtcare, Coventry University, Coventry, United Kingdom

Contributions: Conceptualisation, Methodology, Data collection & analysis, Writing – original draft, Corresponding author

ORCID: <https://orcid.org/0009-0009-0806-0511>

**Dan Edgcumbe**

Affiliation: School of Social Sciences and Humanities, College of Arts and Society, Coventry University, United Kingdom

Contributions: Review and Supervision

ORCID: <https://orcid.org/0000-0002-9748-8019>

**Sally Pezaro**

Affiliation: Department of Healthcare and Communities, Coventry University, Coventry, United Kingdom

Contributions: Review and Supervision

ORCID: <https://orcid.org/0000-0001-5767-0708>

**Ksenija da Silva**

Affiliation: Department of Healthcare and Communities, Coventry University, Coventry, United Kingdom

Contributions: Review and Supervision

ORCID: <https://orcid.org/0000-0002-9232-3143>

**Corresponding Author**

Dr. Sara Arouch

Centre for Intelligent Healthcare, Coventry University

Richard Crossman Building

Coventry, CV1 5RW, United Kingdom
